# Seasonal patterns of SARS-CoV-2 transmission in secondary schools: a modelling study

**DOI:** 10.1101/2022.04.21.22273952

**Authors:** Thi Mui Pham, Ilse Westerhof, Martin C.J. Bootsma, Mirjam E. Kretzschmar, Ganna Rozhnova, Patricia Bruijning-Verhagen

## Abstract

**Background:** The Omicron variant has caused a new wave of SARS-CoV-2 infections worldwide. We explore crucial epidemiological parameters driving seasonal patterns of SARS-CoV-2 transmission in secondary schools and assess various infection control interventions over a 2.5-year time frame.

**Methods:** We developed an agent-based model parameterised with data from secondary schools in the Netherlands. We modelled the circulation of Omicron assuming a stable introduction rate of infections and accounted for uncertainty in epidemiological parameters describing virus transmissibility, susceptibility to reinfection, vaccine immune escape, and waning of sterilising immunity. We quantified the SARS-CoV-2 health burden defined as number of symptomatic student days. We further evaluated the cost-benefit (number of prevented infected students per absent student) for reactive quarantine interventions, regular screening using antigen tests, and annual booster vaccinations.

**Findings:** Durability of sterilising immunity is a key parameter that governs temporal SARS-CoV-2 transmission patterns in secondary schools. Our model predicts pronounced within-school seasonal patterns with dominant autumn outbreaks and smaller winter outbreaks and a maximum prevalence of 2.9% (95% CI: 0.7%-6.6%) symptomatic students during infection peaks. Regular screening and annual booster vaccination may reduce the health burden up to 15% (95% CI: 1.5%-27.8%) and have a higher cost-benefit ratio than reactive quarantine interventions (reduction: 4.3%; 95% CI: -10.1% to 17.6%).

**Interpretation:** Immunity waning will determine the intensity and pattern of SARS-CoV-2 transmission in secondary schools in the medium-term future. If mitigation strategies are needed, screening and annual booster vaccination have the highest cost-benefit by reducing viral transmission with little educational disruption.

## Introduction

To curtail SARS-CoV-2 transmission, many countries implemented school closures or school-based mitigation measures throughout the pandemic.^1,2^ More recently, high population immunity from vaccination and natural infection have allowed easing of mitigation policies in most regions. However, the emergence of the SARS-CoV-2 variant Omicron in November 2021 has significantly hampered the progress in controlling the pandemic. The future epidemic trajectory of SARS-CoV-2 and the need for additional vaccination or temporary re-instalment of mitigation measures remain uncertain.^3^ Mathematical modelling studies have proven very useful in exploring possible scenarios for the medium and long-term trajectory of SARS-CoV-2 and transitioning to an endemic phase at the population level.^4–6^ Already early in the pandemic, a modelling study informed by data on seasonality, immunity and cross-immunity of human coronaviruses suggested that recurrent wintertime outbreaks would probably occur after the initial pandemic wave.^4^ Another study concluded that if SARS-CoV-2 immunological protection is comparable to that of other seasonal human coronaviruses, SARS-CoV-2 will cause no more than a common cold-like disease once the endemic phase is reached.^6^ Keeling and colleagues simulated the potential impact of the Omicron variant in the UK over three months from January to April 2022 under the assumption of lifting control measures.^7^ The authors showed that due to its growth advantage, Omicron can generate high levels of infection that could put a high burden on the healthcare system.

While these studies capture potential future dynamics of SARS-CoV-2 at the population level, longer-term dynamics in specific settings such as secondary schools and the associated burden of infections in school children have not been assessed. Modelling studies in school populations thus far mainly focused on estimating within-school reproduction numbers and on short-term predictions of the potential impact of school-based mitigation measures (e.g., school closure, regular testing, mask-wearing, room ventilation, cohorting) during single waves of the pandemic.^8–10^ Regular screening using rapid antigen tests was found to be efficient in preventing infections while reducing absent student days.^8,10^ Its effectiveness, however, highly depends on testing adherence and turn-around time.^8,9^ A modelling study calibrated to Austrian data found that large infection clusters are effectively prevented by a combination of school-based measures only.^9^

Many of these within-school mitigation interventions take a substantial toll on students’ well-being and education, while infections rarely result in severe morbidity and mortality.^11,12^ It, thus, remains crucial to investigate the expected magnitude of future SARS-CoV-2 transmission in schools, the accompanying burden of infections on students’ health as well as the impact of school-based interventions on virus circulation and school absence. These projections need to account for uncertainties about Omicron’s epidemiological characteristics such as altered transmissibility, disease severity, immune escape, and the rate of immunity waning after vaccination, infection, or both.^13,14^ Here, we developed an agent-based model to investigate the transmission dynamics of SARS-CoV-2 in secondary school children aged 12 to 18 years over a 2.5-year time frame. Our model was parameterised using data from secondary schools in the Netherlands. Starting with the dominance of Omicron in the community at the beginning of 2022, we investigated how uncertainties in important epidemiological characteristics such as intrinsic transmissibility, susceptibility to reinfection, degree of vaccine immune escape, and immunity waning affect the temporal transmission patterns in a secondary school. We further studied the effectiveness and cost-benefit of reactive quarantine interventions, regular screening using rapid antigen tests, and of an annual booster vaccination campaign. Our analyses highlight the key epidemiological parameters for understanding school transmission dynamics and school-based measures that could be effective in a medium-term future.

## Methods

### Data

Field data were collected from a pilot project on rapid antigen testing in secondary schools initiated by the Dutch ministry of education. In brief, between January and April 2021, a representative selection of 45 secondary schools in the Netherlands providing education to 12-18 years-old students in grades 1-6 adopted a risk-based testing policy. Upon report of an index case, school officials identified all school-based contacts of the index case during the presumed infectious period for at least one teaching hour. These students were offered antigen testing on the same day and a repeat test 3-5 days later. Testing was conducted on school premises and performed by a certified test supplier. The invited student and teachers were asked to complete a short questionnaire about COVID-19 symptoms, recent contacts with known infected subjects and details on the number and type of their school contacts. These data, supplemented with data from literature, served as the main input for our model. A detailed description of the pilot is given in Appendix A.

### Agent-based model

We developed an agent-based model to simulate SARS-CoV-2 transmission in a secondary school informed by data previously described. We provide a model overview and most important model parameters in Figure 1 and Table 1, and a brief description below. A more detailed description of the model is given in Appendix B. The code for the model can be found on Github.^15^

**Figure 1.**
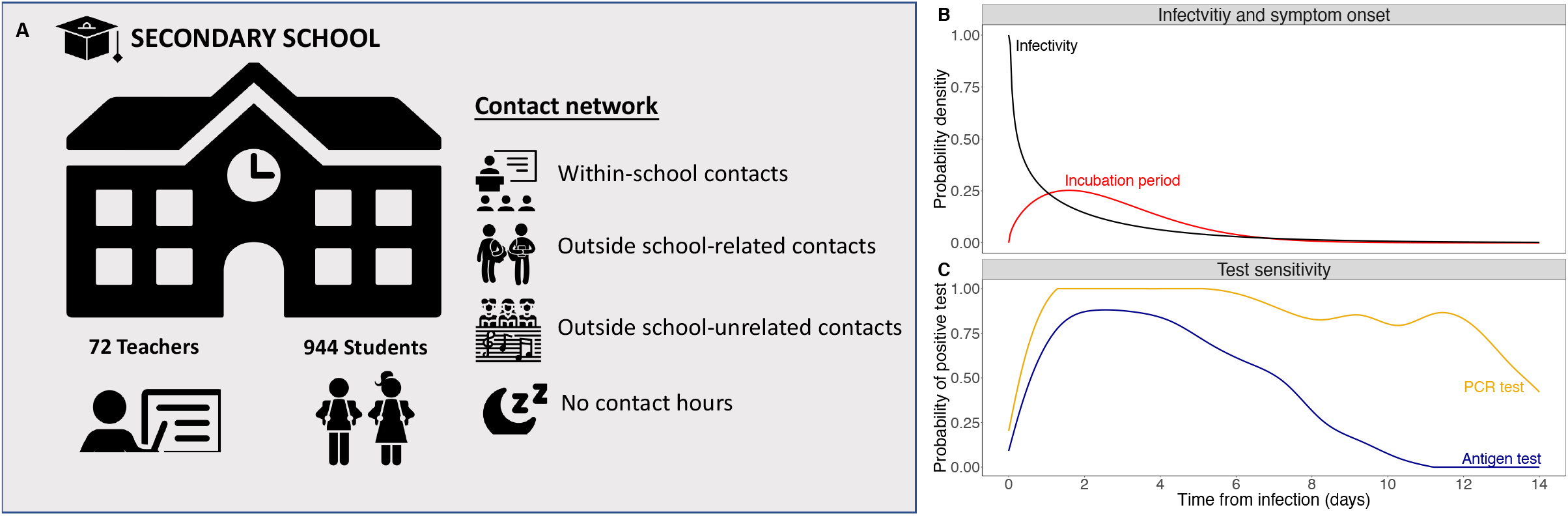
Model overview. (A) Overview of agents and contact network of the agent-based model. (B) Infectivity and symptom onset of infectious individuals. (C) Test sensitivity curve for PCR and antigen test used in the model.

**Table 1.**
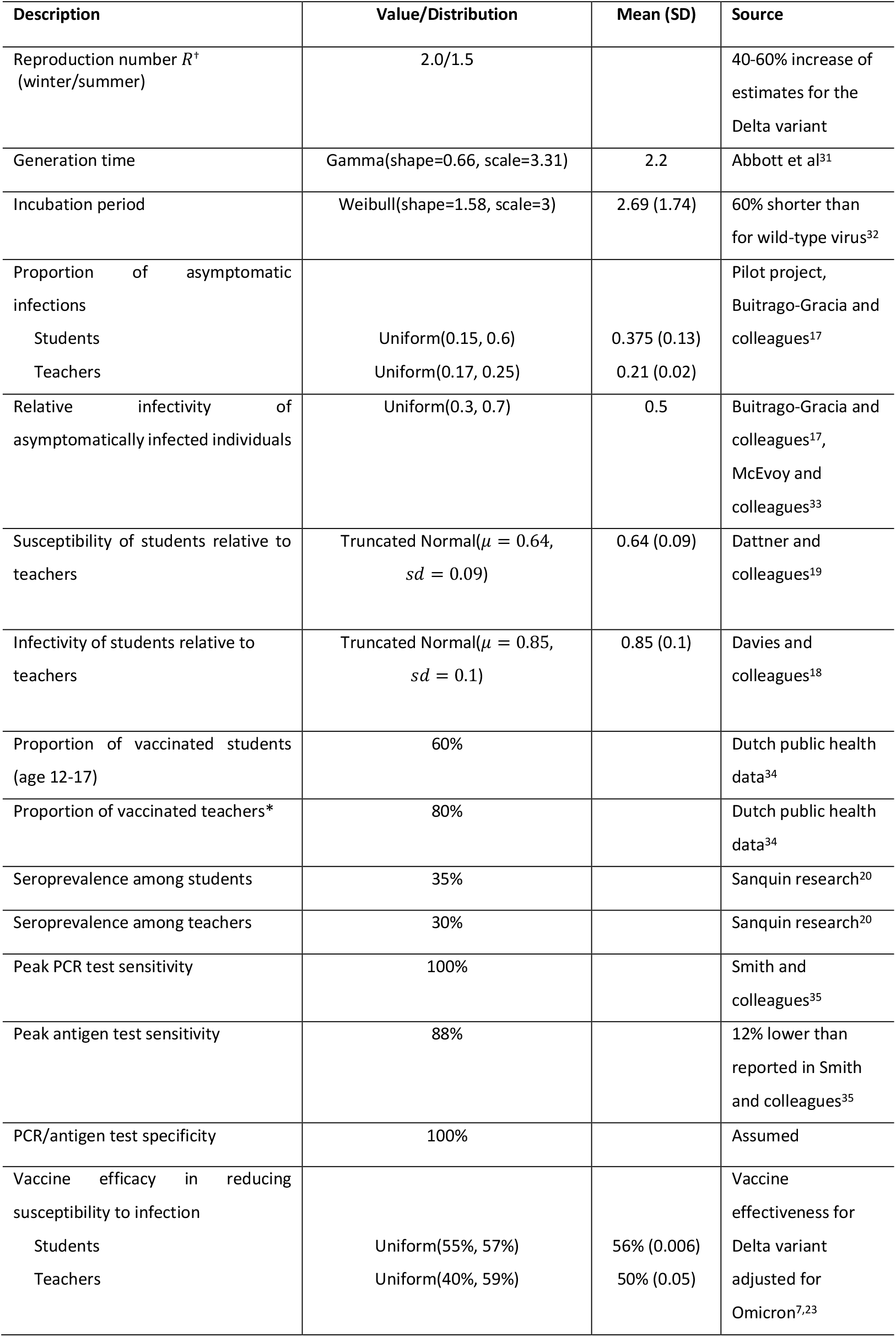
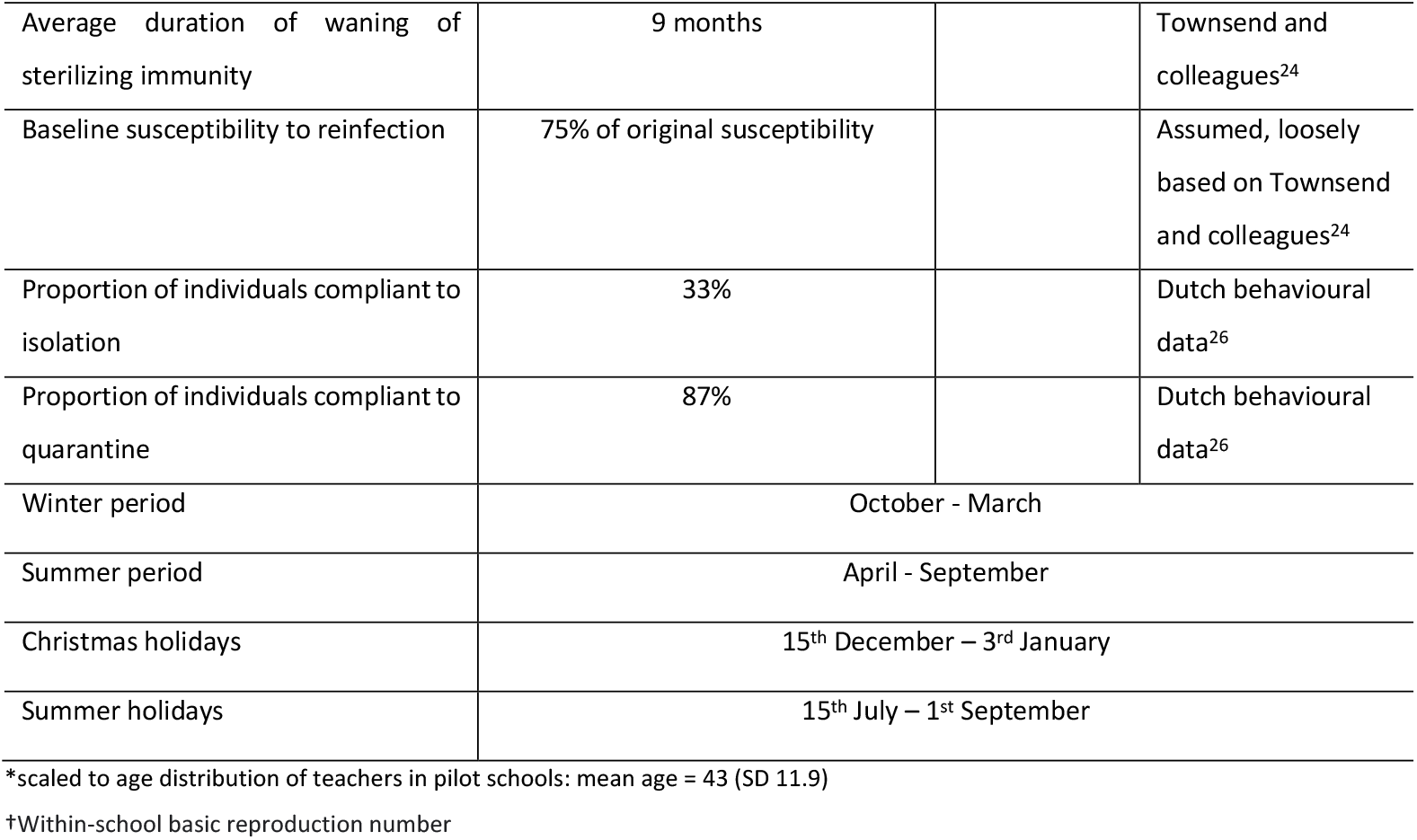
Epidemiological parameters used in the baseline scenario.

We distinguished two types of individuals: (1) students, characterised by the grade and class they belong to; (2) teachers, characterised by the classes they educate. Based on average values reported in the pilot (Appendix A), the secondary school in our model comprises six grades and 944 students in total. Students attend five subjects per day and teachers educate two to three classes per day, resulting in 72 teachers in our model.

#### Contact network

We defined contacts relevant for transmission based on data from the pilot (Appendix A.2). Students reported the number of contacts (defined as conversations at less than 1.5 m and for at least 15 min, or physical touch) with fellow students within the same class and outside their class during and after school hours. Results were summarized into contact matrices in the school environment. We distinguished weekdays (Monday to Friday) and weekends (Saturday and Sunday), and divided the day into three periods of eight hours each, distinguished by the types of contacts:

1. *School hours:* Students and teachers have within-school contacts described by the respective contact matrices (Appendix B).

2. *Outside school hours:* Students are assumed to have two school-related contacts during their leisure time activities. These contacts are randomly sampled from their within-school contacts. Teachers are assumed to have no contacts with other teachers after school hours. Transmission risks caused by school-unrelated contacts are modelled by a constant introduction rate of infected students and teachers (Appenix B).

3. *Night hours:* Neither students nor teachers are assumed to have any contacts during this time.

#### Transmission characteristics

Individuals may be either susceptible, vaccinated, symptomatically infected, asymptomatically infected, or recovered. The baseline infectivity is distributed according a Gamma distribution (mean = 2.2 days) based on results specific for Omicron^16^ and the assumed basic reproduction number. Symptomatically infected individuals are assumed to develop symptoms according to a Weibull-distributed incubation period (mean = 2.7 days). The infectivity per contact is assumed to be on average 50% lower for asymptomatic than for symptomatic individuals,^17^ and additionally 15% lower for students when compared to teachers.^18^ Susceptible individuals may become infected upon direct contact according to the contact network described above but also indirectly through aerosol transmission in a classroom (Appendix B). We assumed age-specific susceptibility to infection based on estimates from studies on the wild-type virus (Table 1).^19^

#### Prior infection

We assumed that 35% of students and 30% of teachers were infected prior to the study period.^20^ Each of those infected individuals is assigned an infection time according to SARS-CoV-2 incidence data from the Netherlands.^21^

#### Vaccination

We used a vaccination coverage of 60% for students and 80% for teachers, and assigned vaccination times according to data from the Netherlands.^22^ Vaccinated teachers are assumed to have received one booster dose while no booster doses were assumed for students. Vaccine efficacies in reducing susceptibility to infection are based on estimates reported for the Delta variant scaled by a factor, representing a lower efficacy for the Omicron variant (Table 1 in Keeling and colleagues).^7,23^ We assumed no direct effect of vaccination on infectivity but rather that it reduces the probability of developing a symptomatic infection, thereby indirectly lowering infectivity.

#### Waning of immunity

We allowed for reinfections after recovery from natural infection or after vaccination. Sterilising immunity wanes exponentially with an average waning time of nine months as estimated by Townsend and colleagues.^24^ In our baseline scenario, we assumed that individuals return to 75% of their original susceptibility value, representing residual protection from previous infection or vaccination. At each reinfection, the probability of a symptomatic infection is assumed to be reduced by 20% in line with reported decrease of COVID-related symptoms after reinfection.^25^

### Simulation scenarios

Simulations were performed over a course of 30 months assuming a start date of 3^rd^ January 2022. We explored the effect of variation of several parameters on the transmission dynamics and on the percentage of students who are (symptomatically) infected, susceptible, and absent due to isolation and (if applicable) quarantine for each week of the study period. We show the mean and 95% uncertainty intervals (95% CI). We defined the health burden on students as the number of symptomatic student days. Since annual booster vaccinations only starts after 1^st^ September 2022, we computed the health burden for the intervention scenarios from that date. As a cost-benefit measure for intervention scenarios, we computed the number of prevented infections per absent student. Fixed parameters including those for the baseline scenario are given in Table 1. Parameters that are varied in other scenarios are given in Table 2.

**Table 2.**
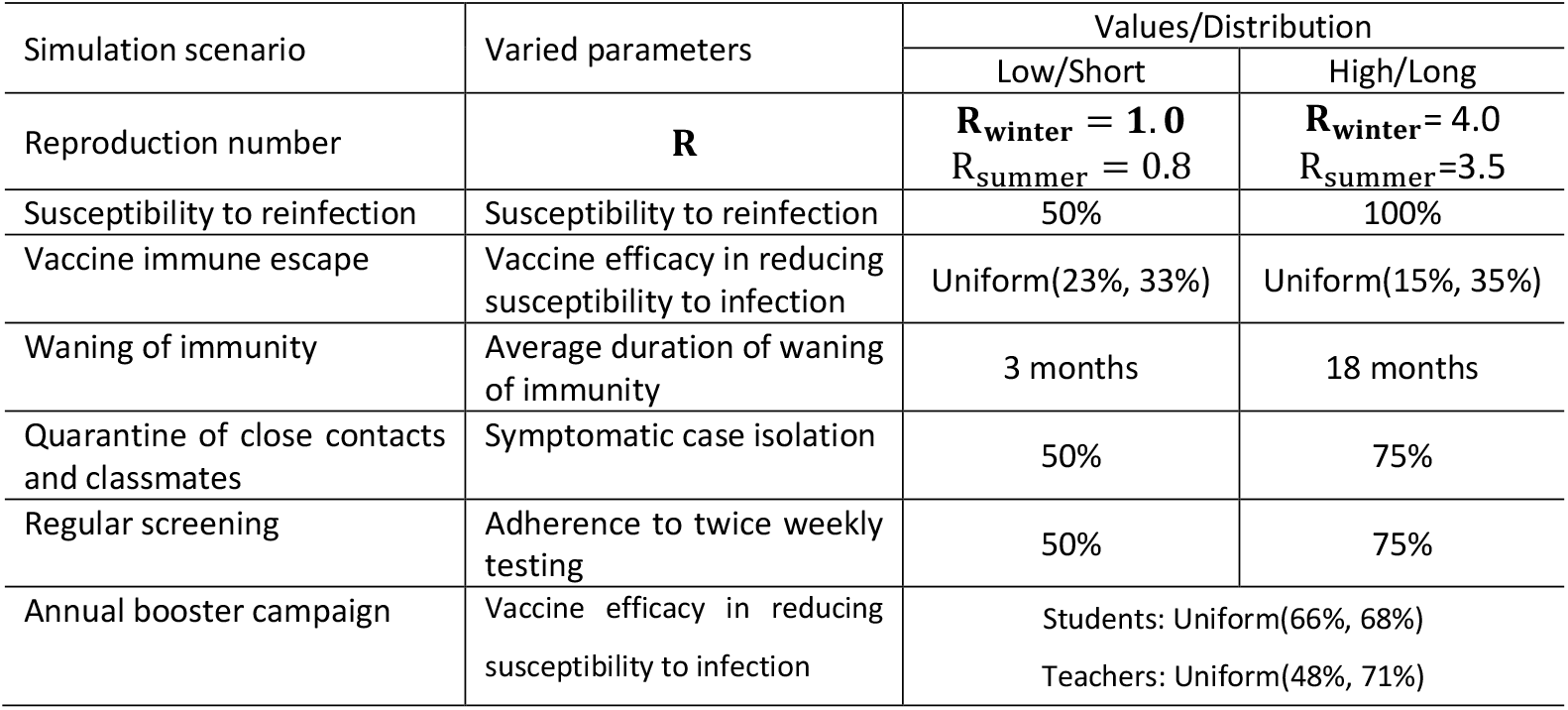
Epidemiological parameters for simulation scenarios.

| Simulation scenario | Varied parameters | Values/Distribution |  |
| --- | --- | --- | --- |
|  |  | Low/Short | High/Long |
| Reproduction number | **R** | **R<sub>winter</sub> = 1.0**<br>R<sub>summer</sub> = 0.8 | **R<sub>winter</sub> = 4.0**<br>R<sub>summer</sub> = 3.5 |
| Susceptibility to reinfection | Susceptibility to reinfection | 50% | 100% |
| Vaccine immune escape | Vaccine efficacy in reducing susceptibility to infection | Uniform(23%, 33%) | Uniform(15%, 35%) |
| Waning of immunity | Average duration of waning of immunity | 3 months | 18 months |
| Quarantine of close contacts and classmates | Symptomatic case isolation | 50% | 75% |
| Regular screening | Adherence to twice weekly testing | 50% | 75% |
| Annual booster campaign | Vaccine efficacy in reducing susceptibility to infection | Students: Uniform(66%, 68%)<br>Teachers: Uniform(48%, 71%) |  |

#### Baseline scenario

The baseline scenario assumes a school-related basic reproduction number for Omicron of 2.0 during the winter period (October to March) and of 1.5 during the summer period (25% decrease compared to winter, April to September), assuming a 40%-60% increase to estimates from the Delta variant.^8,10^ We assumed compliance to isolation guidelines for symptomatically infected students of 33%,^26^ i.e. home isolation for seven days upon a positive PCR test, but no other mitigation measures in schools.

#### Reproduction number

We distinguished two scenarios to account for the uncertainty in the school-related reproduction number: (a) 50% lower reproduction number in winter (*R*_*winter*_ = 1.0, *R*_*summer*_ = 0.75), (b) 100% higher reproduction number in winter (*R*_*winter*_ = 4.0, *R*_*summer*_ = 3.0).

#### Susceptibility to reinfection

We distinguished (a) a lower susceptibility to reinfection of 50% and (b) full susceptibility to reinfection, i.e., 100% of the original susceptibility value.

#### Vaccine immune escape

We assumed (a) 25% lower and (b) 25% higher average vaccine efficacy in reducing susceptibility to reinfection, reflecting higher and lower immune escape in vaccinated individuals, respectively.

#### Waning of immunity

We investigated two alternative average durations of sterilising immunity: (a) 3 months and (b) 18 months, as opposed to 9 months for the baseline scenario.

#### Intervention: Quarantine of close contacts and classmates

Upon a positive test result of a compliant symptomatically infected student, all close contacts and classmates quarantine for ten days. Individuals who have a negative antigen test result on day five after the start of quarantine, may exit quarantine. Assuming an increased case detection in comparison with the baseline scenario, we distinguished (a) 50% and (b) 75% symptomatic case isolation.

#### Intervention: Regular screening

A proportion of students will perform an antigen test twice weekly with (a) 50% and (b) 75% adherence to this screening intervention.

#### Intervention: Booster vaccination

All fully vaccinated students and teachers are assumed to receive one booster dose annually during summer holidays. The efficacy of booster vaccination in reducing the susceptibility to infection is assumed to be increased by 20% with respect to the initial efficacy.

#### Sensitivity analyses

The robustness of our results was tested in sensitivity analyses (Appendix E).

## Results

### Pilot project

Between February and April 2021 (dominance of the Alpha variant), 45 schools with a total of 33,274 students and 3,898 teachers participated for a period of 5 to 12 weeks (mean of 9 weeks). A total of 32 secondary schools (71% of participating schools) reported detailed data on 151 SARS-CoV-2 index cases, resulting in testing of 3652 contacts. The number of index cases reported per school varied between 0 and 9 per week with corresponding community incidence rates varying between 180 and 300 per 100,000 person weeks. Upon the report of each index case, a round of risk-based testing of school contacts was instigated, with one case reported on average per week per school (varying between 0 and 5 per week). Among index cases, 79.5% (n = 120) were students and 20.5% (n = 31) were teachers. The results are summarized in Table 3.

**Table 3.** Results of risk-based testing in schools participating in pilot project in the Netherlands.

|  | Index cases<sup>1</sup><br>No. (%) | Index cases with at least one detected secondary case | Classroom contacts participating in antigen testing<sup>1</sup> | Classroom contacts with positive antigen test | Classroom contacts per index case Mean (SD)<sup>3</sup> |  |
| --- | --- | --- | --- | --- | --- | --- |
|  |  |  |  |  | Close contacts<sup>4</sup> | Non-close contacts |
| Students | 120 (79.5%) | 11 (61.1%) | 2863 (78.7%) | 21 (87.5%) | 1.7 (2.2) | 23.6 (22.6) |
| Teachers | 31 (20.5%) | 7 (38.9%) | 774 (21.2%) | 3 (12.5%) | 2.7 (5.1) | 28.8 (29.3) |
| Total | 151 | 18 | 3637 | 24 | 1.9 (3.0) | 24.4 (23.6) |

<sup>1</sup>Index case: student or teacher with SARS-CoV2 infection reported to the school and identified at testing facility outside school.
<sup>2</sup>Classroom contacts with at least one antigen test performed
<sup>3</sup>Based on the numbers available (58 student index cases and 14 teacher index cases).
<sup>4</sup>Close contacts are defined as individuals who had a contact with the index case <1.5 metres for at least 15 minutes, or a household member of the index case

### Simulation results

#### Baseline scenario

Under our baseline assumptions, we reproduced a large school epidemic after the Christmas holidays at the beginning of 2022 (Appendix Figure 2),^27^ followed by low virus circulation from April till July 2022. Subsequently, large “autumn waves” (September till December 2022/2023) are followed by smaller “winter waves” (January till April 2023/2024) and low virus circulation in spring and summer (April till August 2023/2024, Figure 2A). Due to higher reproduction numbers in winter, we observe higher peaks in autumn than in spring and summer. In our model, a maximum of 26.1% (95% CI: 14.0%-35.5%) of students are infected during the first peak in January 2022, and 8% (95% CI: 3%-15%) of students at the peak of the last autumn wave in 2023. We expect few symptomatic infections and absenteeism after the initial wave in January 2022 with a peak of 2.9% (95% CI: 0.7%-6.6%) of students symptomatically infected and 0.9% (95% CI: 0.1%-1.9%) of students absent from school in mid-October 2022 (Figure 2B-C).

**Figure 2.**
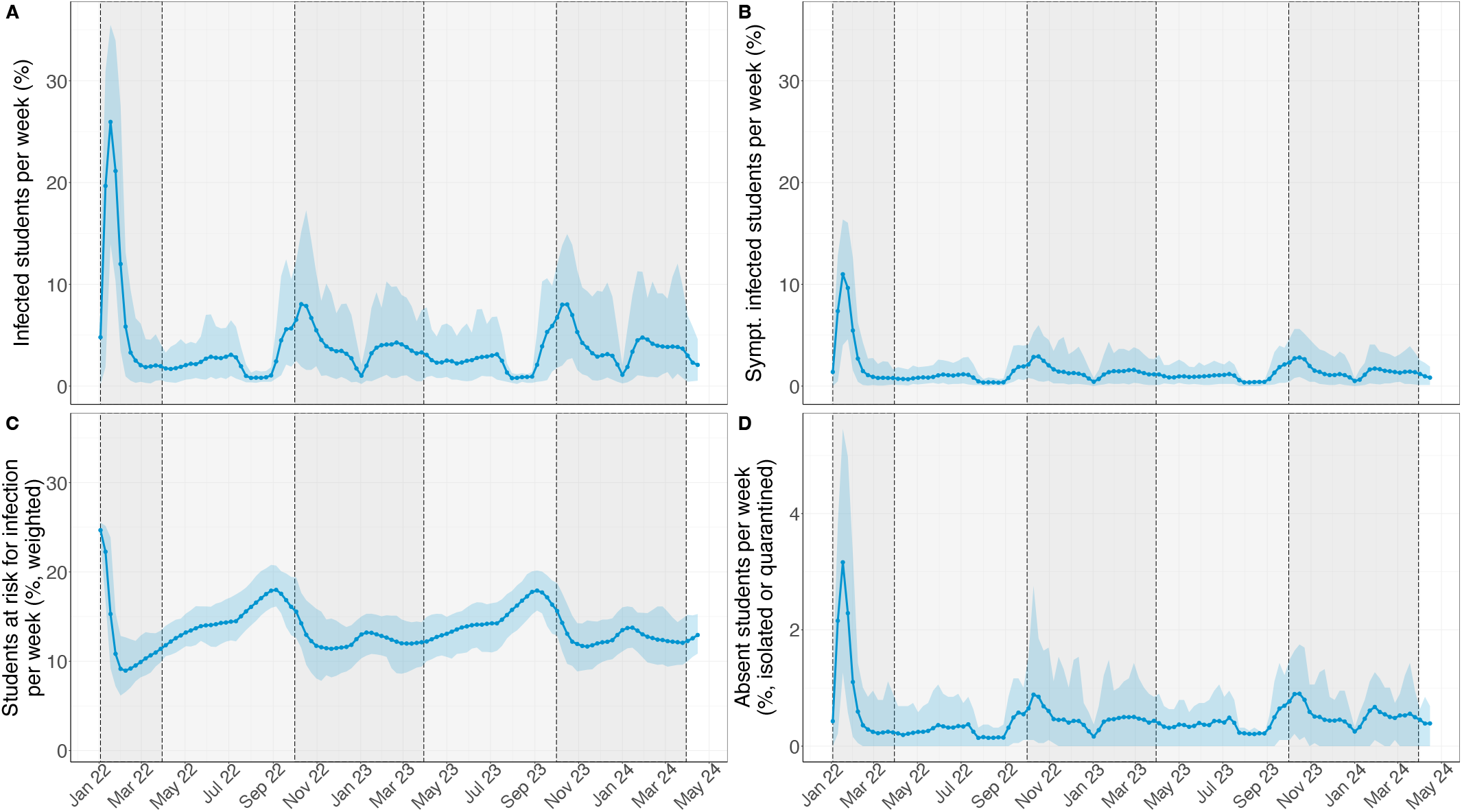
SARS-CoV-2 transmission dynamics for the baseline scenario. Bold points represent the mean value per week over 100 simulations. Shaded coloured areas are 95% uncertainty intervals over 100 simulations. Dark grey background represents the “winter” period (October till March). Light grey background represents the “summer” period (April till September). (A) Proportion of students infected per week due to school-related infections and introductions from community. (B) Proportion of students symptomatically infected per week. (C) Proportion of students at risk for infection per week, weighted by their susceptibility value (with a baseline of 100% for unvaccinated teachers). (D) Proportion of students either isolated or quarantined per week.

#### Reproduction number governs the magnitude and timing of epidemic peaks

An increase in the reproduction number expectedly results in higher peaks of the epidemic waves (Figure 3A), with autumn waves reaching their peaks about a month earlier when compared to the baseline scenario. For lower reproduction numbers, there is only one pronounced initial peak and rather low virus circulation subsequently with only about 2 to 3% of infected students per week.

**Figure 3.**
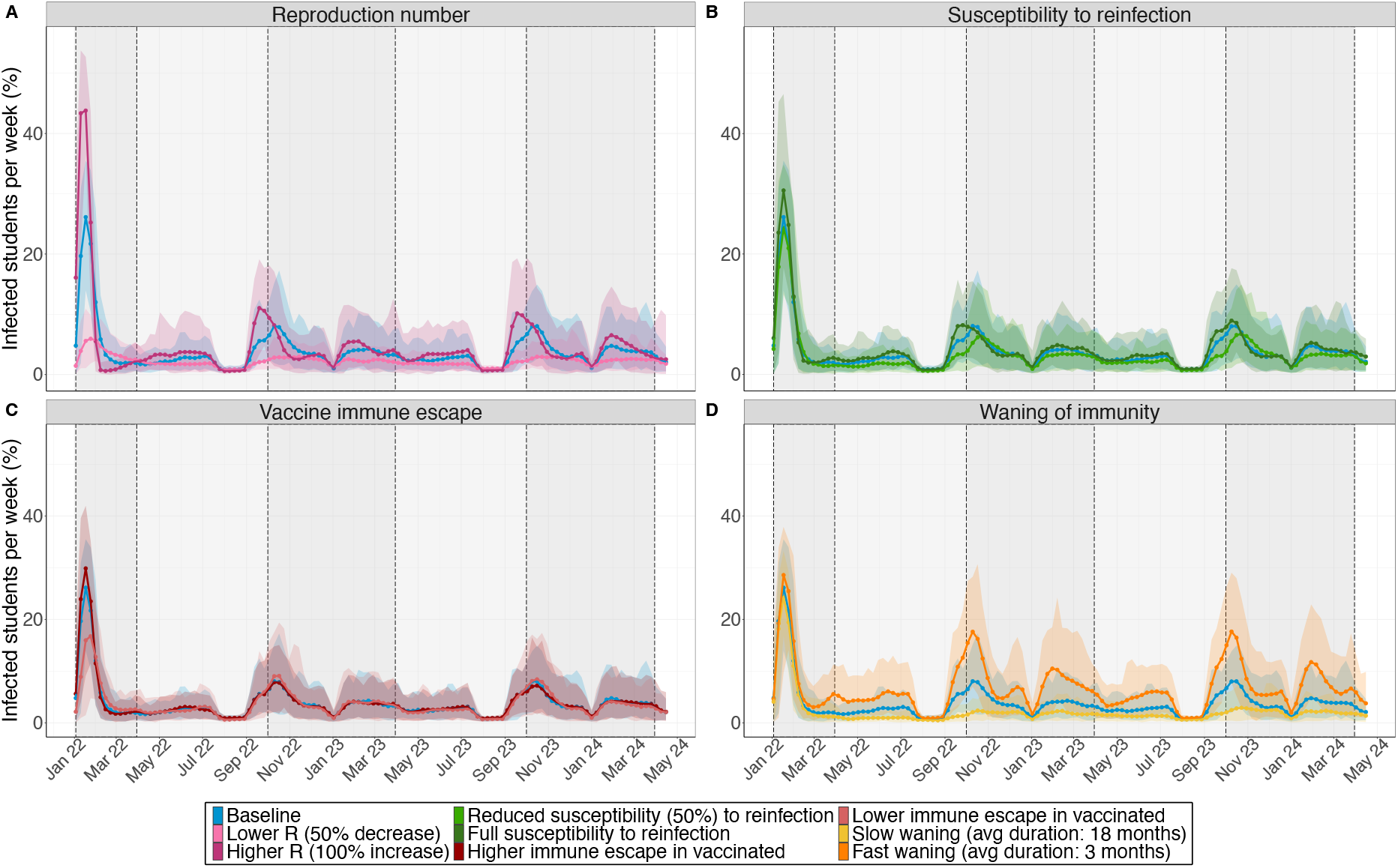
SARS-CoV-2 transmission dynamics for the simulation scenarios. Proportion of students infected with SARS-CoV-2 in each week form 03/01/2022 till 24/05/2024. Bold points represent the mean value per week over 100 simulations. Shaded coloured areas are 95% uncertainty intervals over 100 simulations. Dark grey background represents the “winter” period (October till March). Light grey background represents the “summer” period (April till September). (A) Scenarios where the within-school reproduction number is varied: (i) lower (*R*_*winter*_=1.0, *R*_*summer*_=0.75)and (ii) higher reproduction number (*R*^*wimter*^ =4.0, *R*^summer^ =3.2). (B) Scenarios where susceptibility to reinfection is varied: (i) susceptibility to reinfection is reduced by 50% of the original susceptibility value and (ii) the full susceptibility to reinfection. (C) Scenario with lower vaccine efficacy against susceptibility of infection. (D) Scenarios where average duration of waning of immunity is varied: (i) slow waning (average duration: 18 months) and (ii) fast waning (average duration: 3 months).

#### Small impact of susceptibility to reinfection

Generally, the impact of susceptibility to reinfection seems to be small. If individuals are fully susceptible to reinfection, the peaks of the outbreaks are not only higher but also the wave occurs earlier in comparison to the baseline scenario (Figure 3B). Similarly, if individuals have a lower susceptibility to reinfection, the peaks of the epidemic waves are lower and also slightly delayed (Figure 3B).

#### Varying vaccine immune escape mainly affects the initial peak

A higher virus escape from vaccine-induced immunity leads to a higher initial peak in January 2022 and slightly lower autumn peaks in the next two years. Lower vaccine immune escape causes a smaller initial outbreak but higher peaks in subsequent autumn waves due to the accumulation of susceptible individuals.

#### Durability of sterilising immunity significantly changes infection patterns

If immunity to infection wanes very quickly (on average after three months, Figure 3D), one additional pronounced wave can be expected pre-Christmas from the end of November till mid-December in 2022 and 2023. On the contrary, if immunity wanes slower (on average after eighteen months, Figure 3D), only few infections may be observed after a large initial peak post-Christmas 2021. Among all simulation scenarios waning of immunity has the largest impact on the transmission dynamics and on the health burden (Appendix Figure 4).

#### Impact of interventions

Quarantining classmates and close contacts in reaction to isolation of symptomatically infected students would have had the highest impact in January 2022 (Figure 4A). Its impact is predicted to be small on subsequent outbreaks with larger effects during autumn waves. However, due to lower population immunity, the winter outbreaks are of similar extent when compared to the baseline scenario. Overall, the reduction in health burden of symptomatic student days is small with a mean reduction of 4.3% (95% CI: -10.1% to 17.6%). The proportion of absent students is hugely increased. The cost-benefit of this intervention is low (Figure 5B) throughout the study period with a maximum of 0.32 prevented infected students per absent student.

**Figure 4.**
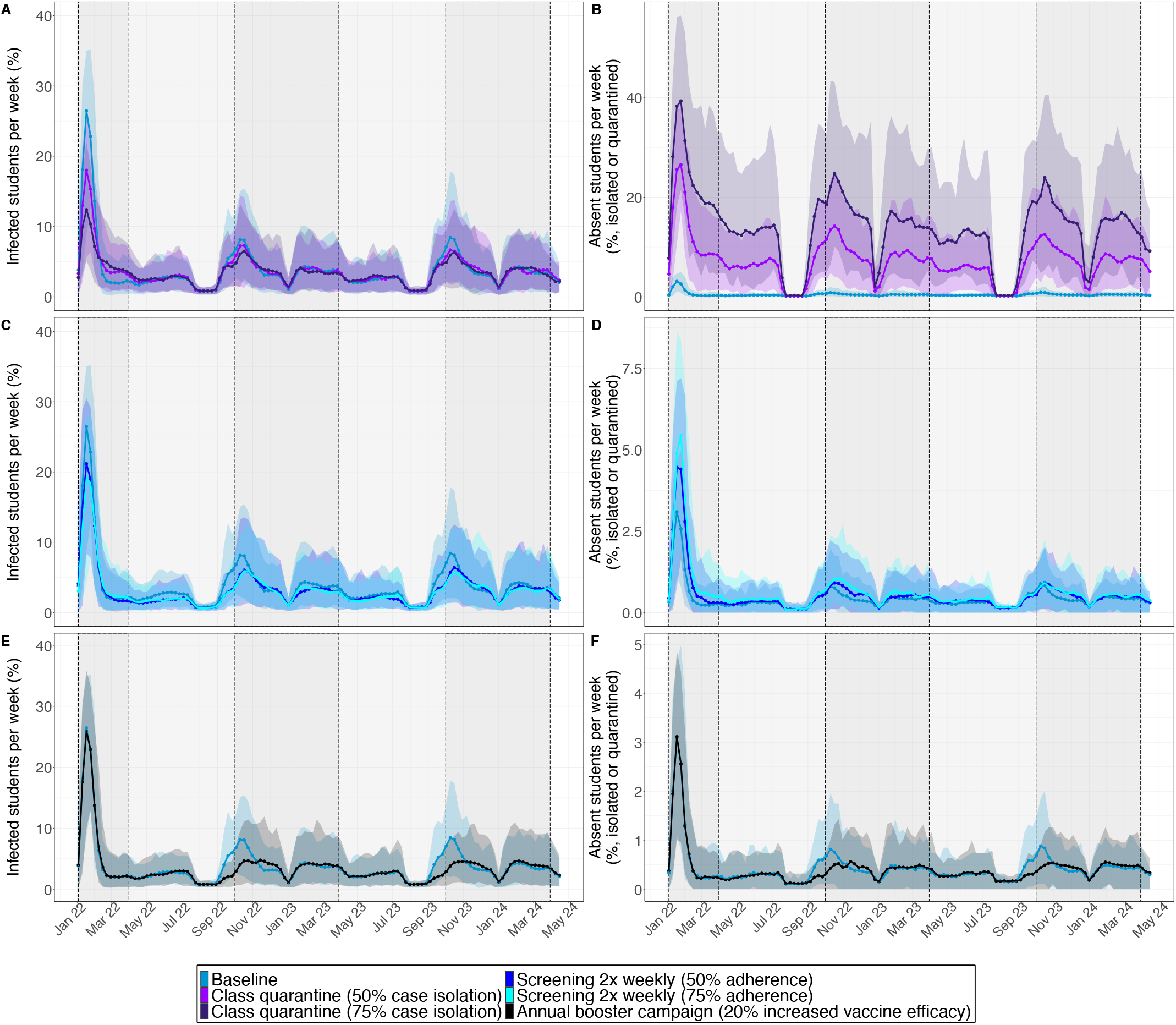
SARS-CoV-2 transmission dynamics for the intervention scenarios. Results are shown for the baseline scenario, quarantine interventions, regular screening and annual booster vaccinations. Bold points represent the mean value per week over 100 simulations. Shaded coloured areas are 95% uncertainty intervals over 100 simulations. Dark grey background represents the “winter” period (October till March). Light grey background represents the “summer” period (April till September). (A), (C), (E) Proportion of students infected with SARS-CoV-2. (B), (D), (F) Proportion of students absent due to isolation of quarantine.

**Figure 5.**
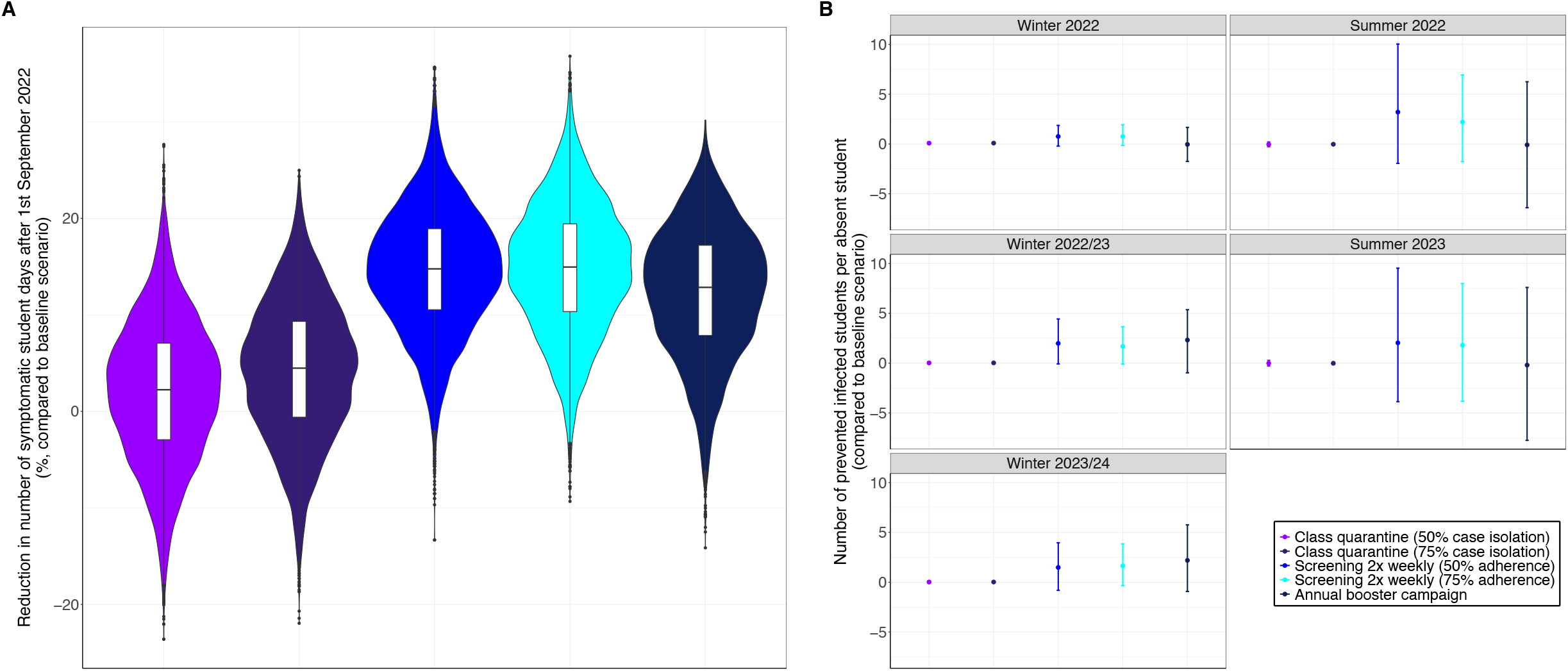
Relative health burden and cost-benefit of intervention scenarios with respect to the baseline scenario. Results are shown for class and close contact quarantine interventions (50% and 75% case isolation), regular screening interventions (50% and 75% adherence), and annual booster vaccinations. (A) Reduction in health burden (defined as the number of symptomatic student days). Since annual booster doses are administered only from September 2022, the reduction is computed for the time period from 01/09/2022 till 24/05/2024. (B) Cost-benefit (number of prevented infected students, compared to baseline, is divided by the mean number of absent students for the respective intervention scenario. The respective numbers are summarised by winter (October till March) and summer period (April till September). All results were obtained by a bootstrapping method. Bold points represent the mean values and error bars represent 95% bootstrapping uncertainty intervals.

Twice weekly screening with 50% and 75% adherence, respectively, prevents at most 5% of infected students (Figure 4C). While the maximum effect is achieved during the initial wave in January 2022 for 75% adherence, screening is more effective than quarantine interventions in preventing infections in subsequent waves. The mean reduction in health burden is 15.1% (95% CI: 1.5% to 27.8%). Screening has the highest cost-benefit of all interventions (Figure 5B). Its mean value is stable over time but its variation strongly depends on the time period. Screening with 50% adherence achieves a maximum of 3.4 (95% CI: -2.0 to 9.9) prevented infected students per absent student in September 2022.

Similarly, annual booster vaccinations are mainly effective in mitigating autumn waves (Figure 4E). Its cost-benefit depends strongly on the time period of the year (Figure 5B), with the highest mean cost-benefit of 2.3 (95% CI: -0.8, 5.3) in winter 2022/23. Due to waning of immunity, the cost-benefit is low in summer periods. Annual booster vaccinations reduce the health burden for students to a similar extent as screening interventions.

## Discussion

Our analyses show that in the baseline scenario where variant characteristics are kept constant during the whole time period of 2.5 years, we expect two distinct epidemic waves during a secondary school year. The highest peaks are expected in autumn due to the accumulation of susceptibles in summer. Peaks are lower in winter than in autumn. This is different to seasonal influenza that usually peaks annually between November and April in the Northern Hemisphere.^28^

Our modelling results suggest that future transmission trajectories in secondary schools are governed by the durability of sterilising immunity. If immunity wanes fast, i.e., on average three months, autumn and winter waves are more pronounced and another wave in spring/summer and one additional peak before the Christmas may be observed. If immunity wanes slow, the number of school-related infections remains low throughout the year.

Our results also show that screening has a higher cost-benefit in terms of number of prevented infections per absent student than reactive quarantine interventions. These results agree with previous results by Colosi and colleagues that assessed the impact of interventions in primary and secondary schools over three months.^8^ Our analyses further show that the cost-benefit of annual booster vaccinations is similar to screening during winter periods. Due to waning of immunity, their cost-benefit is low in summer. Assuming a similar incidence as in October 2021 in the Netherlands, we would expect few symptomatic infections among students even in winter. Hence, interventions might not be needed to reduce the health burden in secondary schools. However, surges in SARS-CoV-2 transmission in the general population may require school-based interventions to mitigate the spread. Our analyses imply that regular screening and annual booster campaigns can help slowing transmissions in schools with little disruption of the education of students.

Qualitatively, our results concur with data from the Netherlands where a large outbreak after the Christmas holidays was reported in adolescents aged 10 to 19 years in 2022.^27^ Our results also agree with previous modelling studies on a population level, which found that due to immunity waning, SARS-CoV-2 will enter regular long-term circulation.^4,5^ Similar to short-term predictions for the effect of potential variants of concern in England by Dyson and colleagues, we found that the magnitude and timings of epidemic peaks highly depend on epidemiological characteristics of the circulating variant.^29^ Our study adds to existing literature by revealing seasonal patterns and highlighting key parameters that characterise SARS-CoV-2 transmission trajectories in secondary schools over a longer time period of 2.5 years. We parameterised our model using data on school characteristics and contact structure in secondary schools in the Netherlands, and accounted for uncertainty in important epidemiological characteristics of the Omicron variant.

Nevertheless, we made several simplifying assumptions in our model. The interpretation of our results has, therefore, limitations. We assumed no further introduction of new variants with substantially different epidemiological characteristics. The timing of such introductions and respective epidemiological characteristics are highly uncertain. Projections on how new variants of concern would impact ongoing transmission dynamics are, therefore, more relevant and feasible on a shorter time scale. Since community incidence on a longer time horizon is difficult to predict, we, further, assumed a constant infection risk for students and teachers from the community throughout the study period. The extent of outbreaks of SARS-CoV-2 infections in schools will depend on the infection risk students and teachers are exposed to outside the school setting. In our model, 33% of symptomatically infected individuals would adhere to self-isolation. These values might vary across different schools and countries, and may impact the predictions on absenteeism and infection prevalence. Finally, in our main analysis, we assumed that vaccine efficacy reducing susceptibility to infection increased by 20% for booster vaccinations. While previous studies have shown an increase in vaccine efficacy against symptomatic disease after booster vaccinations against Omicron,^7^ there is no similar data on efficacy against infection, yet. We performed various sensitivity analyses and showed that our general conclusions remained unaffected.

In conclusion, our results highlight that future transmission trajectories in secondary schools highly depend on the epidemiological characteristics of the circulating SARS-CoV-2 variant. We expect that annual school outbreaks will be dominant in autumn, followed by a smaller winter outbreak. Knowledge about how long immunity against reinfection of SARS-CoV-2 will last in school populations will be decisive for the need of mitigation measures in secondary schools. Of the studied interventions, regular screening or annual booster vaccinations have the most favourable cost-benefit ratio.

## Data Availability

All data produced are available online at: https://github.com/tm-pham/covid19_school_transmission/tree/master/data

## Authors’ contributions

TMP, IW, MCJB, MK, GR and PB developed the conceptual framework of the study. IW and PB accessed and verified all data. IW and TMP analysed the data. TMP developed the code and produced the results of the model. All authors interpreted the results. TMP, GR and PB wrote the article. All authors contributed to and approved the final version of the article.

