## Supplementary material for "Seasonal patterns of SARS-CoV-2 transmission in secondary schools: a modelling study"

**Appendix A. Pilot project data.**

Field data were collected in a pilot project that evaluated a secondary school-based policy of rapid antigen testing. This risk-based testing policy consisted of antigen testing offered to all school contacts of an infected student or teacher who was identified via a positive PCR test outside schools. Between January and April 2021, a representative selection of 45 secondary schools in the Netherlands providing education to 12-18 years-old students in grades 1-6 participated in the pilot project. Participation required schools to adopt a risk-based testing policy and to maintain records of the number and characteristics of index cases, number of contacts invited for risk-based testing and aggregated test results. Upon report of an index case, school officials would identify all school-based contacts who had been in contact with the index case during the presumed infectious period for at least one teaching hour. These students were offered to perform antigen testing on the same day and a repeat test 3 to 5 days later. Testing was conducted on school premises and performed by a certified test supplier. In addition, the invited students and teachers were asked to complete a short questionnaire about COVID-19 symptoms, recent contacts with known infected subjects and details on the number and type of their school contacts. The pilot project yielded detailed data from 32 distinct schools on 151 SARS-CoV-2 infections and 3652 tested contacts. These data, supplemented with data from literature, served as the main input for the agent-based transmission model. The detailed results of the pilot project are provided in Appendix Tables 1-13.

### A.1 General school characteristics from the pilot data.

**Appendix Table 1. Pilot data - School characteristics (Median and IQR).**

| Grade | Number of classes per grade | Number of students per grade |
|---|---|---|
| 1 | 7 (3.5-9) | 158 (73-223.5) |
| 2 | 6 (4-10.5) | 164 (67.5-241) |
| 3 | 8 (4-9.5) | 179 (59-238.5) |
| 4 | 7 (4.5-11.5) | 144 (68-294) |
| 5 | 5 (3-7.5) | 129 (58.5-206.5) |
| 6 | 2 (2-3) | 62 (37-78) |

**Appendix Table 2. Pilot data - Overview of teachers and staff in schools.**

| | Median (IQR) |
|---|---|
| Number of teachers | 77 (35.5-129.5) |
| Number of other staff | 18 (0-27.5) |

**Appendix Table 3. Overview of occupancy of students and teachers in secondary schools since 1 March 2021.** Based on data available from 12 schools; schools with special and practical education excluded.

| | Mean (SD) | Median (IQR) |
|---|---|---|
| Number of students per day in school (per 1000 students) | 1131.6 (616.7) | 988.5 (715-1424) |
| Number of teachers at school per week (per 1000 students) | 104.8 (23.8) | 101.5 (85-117.5) |
| Number of teachers at school per day (per 1000 students) | 17.9 (16.7) | 21 (0-29.5) |
| Number of grades a teacher educates per day | 30.2 (24.3) | 23 (14-40) |
| Number of classes educated by a teacher per day | 149 (98.9) | 150 (90-200) |
| Number of students educated by a teacher per day | 1131.6 (616.7) | 988.5 (715-1424) |

**Appendix Table 4. Occupancy stratified by grade since 1 March 2021.** Based on data available from 12 schools; schools with special and practical education excluded.

| | | **Average number of days of education in school** | | **Average number of hours present at school per school day** | | **Average number of teachers a student is educated by at school per school day** | |
|---|---|---|---|---|---|---|---|
| **Grade**† | **Age** | Mean (SD) | Median (IQR) | Mean (SD) | Median (IQR) | Mean (SD) | Median (IQR) |
| Grade 1 | 12 | 2.3 (0.7) | 2.5 (2-2.5) | 5.2 (1.3) | 5 (4.5-6) | 4.7 (1.3) | 5 (3.5-5) |
| Grade 2 | 13 | 2.5 (0.9) | 2.5 (2-2.5) | 5.2 (1.3) | 5 (4.5-6) | 4.8 (1.3) | 5 (4.5-6) |

| | | | | | | | |
|---|---|---|---|---|---|---|---|
| Grade 3 | 14 | 2.5 (0.9) | 2.5 (2-2.5) | 5.1 (1.1) | 5 (4.5-6) | 4.7 (1.3) | 5 (3.5-5) |
| Grade 4 - Exam year | 15 | 4.0 (1.3) | 5 (2.5-5) | 5.5 (1.2) | 5 (5-6.5) | 4.9 (1.4) | 5 (4-6) |
| Grade 4 - No Exam year | 15 | 2.2 (0.3) | 2.5 (2-2.5) | 5.2 (1.1) | 5 (5-6) | 5.1 (1.1) | 5 (5-5) |
| Grade 5 - Exam year | 16 | 3.9 (1.3) | 5 (2.5-5) | 5.3 (1.2) | 5 (5-6) | 5.2 (1.2) | 5 (4.5-6) |
| Grade 5 - No Exam year | 16 | 2.2 (0.4) | 2.5 (2-2.5) | 5.3 (1.2) | 5 (5-6) | 5.1 (1.2) | 5 (4.5-5.5) |
| Grade 6 - Exam year | 17 | 4.1 (1.2) | 5 (3-5) | 5.3 (1.2) | 5 (5-6) | 5.2 (1.2) | 5 (4.5-6) |

† Number of grades differs per level.

### A.2 Results on index cases and secondary cases

**Appendix Table 5. Overview of participating schools, antigen testing via pilot and the number of secondary cases.**

| **Week** | **Number of participating schools** | **Number of students in participating schools** Mean (SD) | **Number of schools with testing** | **Average number of students in schools with testing** Mean (SD) | **Number of schools with positive cases** | **Average number of students in schools with positive cases** Mean (SD) | **Number of index cases with testing** | **Number of index cases with positive cases** | **Number of positive cases (range per index)** |
|---|---|---|---|---|---|---|---|---|---|
| 1/3/2021 | 29 | 601.4 (445.6) | 2 | 1329 (172.5) | 2 (100%) | 1329 (172.5) | 2 | 2 (100%) | 5 (1-4) |
| 8/3/2021 | 33 | 623.1 (497.5) | 5 | 789.8 (542.7) | 1 (20%) | 438 | 6 | 1 (16.7%) | 2 |
| 15/3/2021 | 46 | 768.8 (562.3) | 19 | 1011.8 (574) | 3 (15.8%) | 903.7 (254.1) | 34 | 5 (14.7%) | 7 (1-2) |
| 22/3/2021 | 46 | 768.8 (562.3) | 17 | 962.5 (636.4) | 3 (17.6%) | 927 (1103.8) | 32 | 3 (9.4%) | 3 (1) |
| 29/3/2021 | 46 | 768.8 (562.3) | 18 | 1092.9 (608.7) | 2 (11.1%) | 1168 (55.2) | 21 | 2 (9.5%) | 3 (1) |
| 5/4/2021 | 46 | 768.8 (562.3) | 12 | 937.3 (519.8) | 0 (0.0%) | 0 | 16 | 0 (0.0%) | 0 |
| 12/4/2021 | 45 | 780.1 (564.7) | 18 | 1096.3 (574.9) | 2 (11.1) | 950.5 (330.2) | 30 | 2 (6.7%) | 2 (1) |
| 19/4/2021 | 45 | 780.1 (564.7) | 11 | 1019.9 (502.5) | 2 (18.2%) | 1156.5 (38.9) | 12 | 2 (16.7%) | 2 (1) |

**Appendix Table 6. Index cases notified in the pilot project under risk-based testing policy.** The results are based on data available from 7 schools and scaled to 1000 students per school.

| Week | Students | | Teachers | |
|---|---|---|---|---|
| | Mean (SD) | Median (IQR) | Mean (SD) | Median (IQR) |
| All period | 14 (23.6) | 7.5 (1-11.5) | 2.6 (5.8) | 0 (1.5-7) |
| 1/3/2021 | 2.3 (6) | 0 (0-0) | 0 (0) | 0 (0-0) |
| 8/3/2021 | 4.6 (6.2) | 0.5 (0-8) | 0 (0) | 0 (0-0) |
| 15/3/2021 | 0.7 (1.3) | 0 (0-1) | 0.9 (2.4) | 0 (0-0) |
| 22/3/2021 | 2.7 (4.8) | 0 (0-3) | 1.4 (3.6) | 0 (0-0) |
| 29/3/2021 | 1.9 (4.7) | 0 (0-0.5) | 0.3 (0.5) | 0 (0-0.5) |
| 5/4/2021 | 0.3 (0.7) | 0 (0-0) | 0 (0) | 0 (0-0) |
| 12/4/2021 | 1.6 (2.7) | 0 (0-2.5) | 0.1 (0.2) | 0 (0-0) |
| 19/4/2021 | 0.9 (0.2) | 0 (0-0) | 0 (0) | 0 (0-0) |

**Appendix Table 7. Number of participants in rapid antigen testing rounds in the pilot project.** The results are based on data available for 84 index cases and testing in 24 schools. 3156 contacts were exposed and eligible for testing. Every contact was asked to get tested both in test round 1 (same or following day as exposure) and test round 2 (3-5 days after test round 1).

| | Participants | | Number of contacts tested per index case and test round | |
|---|---|---|---|---|
| | Exposed | Tested | Mean (SD) | Median (IQR) |
| Test round 1 (same day) | 3156 | 1426 (45.2%) | 56.3% (28%) | 53.3% (31.6-78.0) |
| Test round 2 (3-5 days later) | 3156 | 1165 (36.9%) | 47.1 (28.3%) | 44.4% (25.8-62.8%) |
| Participated in at least in one test round | 3156 | 1621 (51.4%) | 61.5% (27.2) | 58.2 (43.4-90.4%) |

**Appendix Table 8. Secondary attack rate stratified by teacher and student.** Secondary attack rate is defined as the number of index cases with at least one secondary case divided by the number of index cases. This includes the index cases for which antigen rapid testing was performed.

| | Total number of index cases | Index cases with secondary transmission | Index cases without secondary transmission | Secondary attack rate | CI 95% |
|---|---|---|---|---|---|
| Teachers | 31 | 7 | 24 | 22.60% | 10.3-41.5 |
| Students | 120 | 11 | 108 | 9.20% | 4.9-16.2 |

**Appendix Table 9. Transmission rate stratified by student and teacher.** Transmission rate is defined as the number of secondary cases divided by the number of participants.

| | Total tested | Secondary cases | Not infected after exposure | Transmission rate | CI 95% |
|---|---|---|---|---|---|
| Teachers | 762 | 3 | 759 | 0.39% | 0.1-1.2 |
| Students | 2813 | 21 | 2792 | 0.75% | 0.48-1.17 |

**Appendix Table 10. Proportion of symptomatic infections among secondary cases.**

| Symptoms† | All | Students | Teachers |
|---|---|---|---|
| - Symptomatic infection | 18 (75.0%) | 15 (71.4%) | 3 (100%) |
| - Pre-symptomatic | 3 (12.5%) | 3 (14.3%) | 0 (0.0%) |
| - Asymptomatic infection | 3 (12.5%) | 3 (14.3%) | 0 (0.0%) |
| † Symptomatic: symptoms during antigen application; Pre-symptomatic: symptom onset in week after antigen test; Asymptomatic: no symptoms. | | | |

### A.2 Student and teacher contact networks.

**Appendix Table 11. Questionnaire for participants regarding their contact network.** The questionnaire was sent to teachers and students who were part of the testing population (students and teachers who were in the same classroom for at least one teaching hour and gave consent for pilot). The original questions were provided in Dutch and translated into English.

| | **Questions translated into English** | **Questionnaire** |
|---|---|---|
| 1 | Which grades do you educate? | Teachers |
| 2 | How many students in this school do you educate in total? | Teachers |
| 3 | With how many students do you have class for all subjects together? | Students |
| 4 | How many of your classmates did you chat with or touch yesterday or the previous day at school? (proxy for close contact) | Students |
| 5 | How many other students (other than classmates) did you chat with or touch yesterday or the previous day at school? | Students |
| 6 | How many of your classmates did you chat with or touched outside of school yesterday? Note; If today is Monday, enter the number you interacted with or touched on Saturday and Sunday? | Students |
| 6.1 | How many of them do you meet at least once a week outside of school? | Students |
| 7 | How many of your schoolmates with whom you are not in class did you chat or touch outside of school yesterday? Note; If today is Monday, enter the number you interacted with or touched on Saturday and Sunday? | Students |
| 7.1 | How many of them do you meet at least once a week outside of school? | Students |
| 8 | How many students that you educated did you chat with or touch yesterday or the previous day at school? | Teachers |
| 9 | How many students did you not teach, chat with or touch yesterday or the previous day at school? | Teachers |
| 10 | How many colleagues did you chat with or touch yesterday or the previous day at school? | Teachers |
| 11 | How many colleagues did you chat with or touch yesterday outside working hours? Note; If today is Monday, enter the number you interacted with or touched on Saturday and Sunday? | Teachers |
| 11.1 | How many of them do you meet at least once a week outside school? | Teachers |
| 12 | How many other people (excluding family members) did you chat with or touch yesterday? Note; If today is Monday, enter the number you interacted with or touched on Saturday and Sunday? | Teachers and students |
| 12.1 | How many of them do you meet at least once a week? | Teachers and students |
| 13 | In the two weeks prior to the test, did you have a close contact with a person with a confirmed coronavirus infection outside school? | Teachers and students |
| 14 | Did you participate in group sports and / or activities in the past two weeks? | Teachers and students |
| 15 | In the past two weeks, how often did you stay indoors with non-family members in one room for longer than 15 minutes (excluding shops and school)? | Teachers and students |
| 16 | In the week prior to the test, where did you contact the infected person for whom you are taking this test? | Teachers and students |

**Appendix Table 12. Questionnaire for schools regarding contact network.** The questionnaire was offered at the completion of the pilot project. The original questions were provided in Dutch and translated into English.

| | **Questions translated into English** |
|---|---|
| 1 | How many students were educated at school at least 1 day a week in the period from 1 March to 24 April? |
| 2 | How many students have been at school per day on average since 1 March? |
| 3 | What is the average number of days of education at school per grade per week since 1 March? |
| 4 | What is the average number of hours a student is present at school per grade since 1 March? |
| 5 | What is the average number of teachers that give lessons to students, per student at school per day per grade since March 1? |
| 6 | How many teachers taught at school at least 1 day a week in the period from 1 March to 24 April? |
| 7 | How many teachers have there been on average per day at school since 1 March? |
| 8 | How many classes does a teacher give on an average education per day at school since 1 March? |
| 9 | How many students does a teacher educate per day at school since 1 March? |

| 10 | How many different grades does a teacher educate per day at school since 1 March? |
|---|---|

**Appendix Table 13. Contacts outside school. The questionnaire completed by 2867 students and 663 teachers.** A contact was defined as a person with whom the participant had a conversation with (at less than 1.5 m distance and for at least 15 min) or physically touched.

| | | **Number of contacts they met the day before/weekend outside school†** | | **Number of these contacts they meet at least twice a week†** | |
|---|---|---|---|---|---|
| | | Mean (SD) | Median (IQR) | Mean (SD) | Median (IQR) |
| Students | Contact with classmates | 2.1 (3.7) | 1 (0-3) | 1.1 (2.4) | 0 (0-1) |
| | Contacts with other students (excl. classmates) | 1.4 (3.7) | 0 (0-2) | 0.8 (2.2) | 0 (0-1) |
| | Other contacts (excl. household members) | 3.6 (5.9) | 2 (0-5) | 2 (4.7) | 0 (0-2) |
| Teachers | Contact with colleagues | 0.5 (1.4) | 0 (0-0) | 0.1 (0.4) | 0 (0-0) |
| | Other contacts (excl. household members) | 3.3 (4.9) | 2 (0-4) | 1.3 (3.3) | 0 (0-2) |

† Contacts at the weekend reported on Monday were divided by 2 for correction.

**Appendix Table 14. Participant contact with index case.**

| **Contact with index** | **All participants** | **Secondary cases** | **Not infected after exposure** |
|---|---|---|---|
| Seated within 1.5 meter from index | 196 (14.2%) | 6 (33.3%) | 190 (13.9%) |
| Outside school | 20 (1.4%) | 1 (5.6%) | 19 (1.4%) |
| During lunch break | 102 (7.4%) | 3 (16.7%) | 99 (7.2%) |
| In same class | 760 (54.9%) | 4 (22.2%) | 756 (55.3%) |
| Other | 70 (5.1%) | 1 (5.6%) | 69 (5.0%) |
| Unknown | 237 (17.1%) | 3 (16.7%) | 234 (17.1%) |

**Appendix Table 15. Contacts of teachers and staff.**

| | Mean (SD) | Median (IQR) |
|---|---|---|
| Number of contacts of teachers with students educated last school day | 19.6 (27.4) | 10 (3-25) |
| Number of contacts of teachers with students not educated last school day | 8.4 (21.1) | 3 (0-10) |
| Number of contacts of teachers with colleagues last work/school day | 8.3 (7.1) | 6.5 (4-10) |

**Appendix Table 16. Number of student contacts that had to go into quarantine because of close contact.**

| | **Mean (SD)** |
|---|---|
| Number classmates | 1.7 (2.4) |
| Number of other persons (excl. class mates) | 0.4 (1.3) |

**Appendix Table 17. Contact matrix of students having contact with students.**

| | | Contacts with students within the same class | | Contacts with students from other classes/grades | | | | | | | | | | | |
|---|---|---|---|---|---|---|---|---|---|---|---|---|---|---|---|
| | | | | Grade 1 | | Grade 2 | | Grade 3 | | Grade 4 | | Grade 5 | | Grade 6 | |
| | | Mean (SD) | Median (IQR) | Mean (SD) | Median (IQR) | Mean (SD) | Median (IQR) | Mean (SD) | Median (IQR) | Mean (SD) | Median (IQR) | Mean (SD) | Median (IQR) | Mean (SD) | Median (IQR) |
| Respondents per grade | 1 | 5.5 (3.8) | 5.0 (3-7) | 3.1 (5.4) | 1 (0-3.0) | 0.20 (0.77) | 0 (0-0) | 0.13 (0.43) | 0 (0-0.0) | 0.07 (0.29) | 0 (0-0) | 0.05 (0.25) | 0 (0-0) | 0.03 (0.22) | 0 (0-0.0) |
| | 2 | 5.2 (3.9) | 4.0 (3-7) | 2.37 (6.46) | 0 (0-0.5) | 4.69 (5.95) | 2 (0-6) | 0.81 (2.52) | 0 (0-0.5) | 0.47 (2.31) | 0 (0-0) | 0.31 (2.31) | 0 (0-0) | 0.32 (2.34) | 0 (0-0.0) |
| | 3 | 5.6 (4.1) | 5.0 (3-8) | 0.60 (3.10) | 0 (0-0.0) | 0.79 (3.33) | 0 (0-0) | 4.66 (5.68) | 3 (1-6.0) | 0.32 (0.84) | 0 (0-0) | 0.04 (0.32) | 0 (0-0) | 0.05 (0.34) | 0 (0-0.0) |
| | 4 | 8.1 (8.0) | 6.0 (3-10) | 1.42 (5.82) | 0 (0-0.0) | 1.32 (5.60) | 0 (0-0) | 1.76 (5.79) | 0 (0-1.0) | 7.03 (7.71) | 5 (2-10) | 1.22 (3.45) | 0 (0-1) | 0.54 (2.97) | 0 (0-0.0) |
| | 5 | 8.3 (7.7) | 6.0 (4-10) | 0.09 (0.64) | 0 (0-0.0) | 0.08 (0.37) | 0 (0-0) | 0.09 (0.32) | 0 (0-0.0) | 0.95 (2.56) | 0 (0-1) | 7.05 (9.09) | 5 (2-10) | 0.78 (1.80) | 0 (0-1.0) |
| | 6 | 10.6 (8.8) | 8.5 (5-15) | 0.42 (3.18) | 0 (0-0.0) | 0.43 (3.09) | 0 (0-0) | 0.43 (3.12) | 0 (0-0.0) | 0.71 (3.63) | 0 (0-0) | 1.49 (3.77) | 0 (0-1) | 6.78 (7.13) | 5 (2-10.0) |

**Appendix Table 18. Measures in secondary schools in the Netherlands during 2020-2021. The shading shows the holidays. Note; the Netherlands contains of three regions with different weeks of summer holidays. Light shading shows the weeks with summer holidays for some regions and dark shading shows the weeks that all schools have holidays.**

| | 2020 (weeks) | | | | | | | | | | | | | | | | | | | | | | | | | | | | | | | | | | | | | | | | | | | | | | | | | | | | |
|---|---|---|---|---|---|---|---|---|---|---|---|---|---|---|---|---|---|---|---|---|---|---|---|---|---|---|---|---|---|---|---|---|---|---|---|---|---|---|---|---|---|---|---|---|---|---|---|---|---|---|---|---|---|
| | 1 | 2 | 3 | 4 | 5 | 6 | 7 | 8 | 9 | 10 | 11 | 12 | 13 | 14 | 15 | 16 | 17 | 18 | 19 | 20 | 21 | 22 | 23 | 24 | 25 | 26 | 27 | 28 | 29 | 30 | 31 | 32 | 33 | 34 | 35 | 36 | 37 | 38 | 39 | 40 | 41 | 42 | 43 | 44 | 45 | 46 | 47 | 48 | 49 | 50 | 51 | 52 | 53 |
| Closure/digital education | | | | | | | | | | | | X | X | X | X | X | X | X | X | X | X | X | | | | | | | | | | | | | | | | | | | | | | | | | | | | | X | | |
| Half-occupancy (Hybrid education - 1/4 in-school) | | | | | | | | | | | | | | | | | | | | | | | X | X | X | X | X | | | | | | | | | | | | | | | | | | | | | | | | | | |
| Full in person | | | | | | | | | | | | | | | | | | | | | | | | | | | | X | X | | | | | X | X | X | X | X | X | X | X | X | X | X | X | X | X | X | X | X | | | |
| 1.5 m distance to classmates | | | | | | | | | | | | | | | | | | | | | | | X | X | X | X | X | | | | | | | | | | | | | | | | | | | | | | | | | | |
| 1.5 m distance to teachers | | | | | | | | | | | | | | | | | | | | | | | | | | | | X | X | | | | | X | X | X | X | X | X | X | X | X | X | X | X | X | X | X | X | X | | | |
| Antigen testing (2x pw preventive/risk-based) | | | | | | | | | | | | | | | | | | | | | | | | | | | | | | | | | | | | | | | | | | | | | | | | | | | | | |
| Masking | | | | | | | | | | | | | | | | | | | | | | | | | | | | | | | | | | | | | | | | | | | | | | | | | X | X | | | |
| Symptomatic persons quarantined | | | | | | | | | | | | | | | | | | | | | | | X | X | X | X | X | X | X | | | | | X | X | X | X | X | X | X | X | X | X | X | X | X | X | X | X | X | | | |
| Quarantine class (>2cases) | | | | | | | | | | | | | | | | | | | | | | | | | | | | | | | | | | | | | | | | | | | | | | | | | | | | | |
| Holidays | | | | | | | | | | | | | | | | | | | | | | | | | | | | * | * | X | X | X | X | * | * | | | | | | | | | | | | | | | | | X | X |
| *summer holidays differ per region. | | | | | | | | | | | | | | | | | | | | | | | | | | | | | | | | | | | | | | | | | | | | | | | | | | | | | |

| | 2021 (weeks) | | | | | | | | | | | | | | | | | | | | | | | | | | | | | | | | | | | | | | | | | | | | | | | | | | | | |
|---|---|---|---|---|---|---|---|---|---|---|---|---|---|---|---|---|---|---|---|---|---|---|---|---|---|---|---|---|---|---|---|---|---|---|---|---|---|---|---|---|---|---|---|---|---|---|---|---|---|---|---|---|---|
| | 1 | 2 | 3 | 4 | 5 | 6 | 7 | 8 | 9 | 10 | 11 | 12 | 13 | 14 | 15 | 16 | 17 | 18 | 19 | 20 | 21 | 22 | 23 | 24 | 25 | 26 | 27 | 28 | 29 | 30 | 31 | 32 | 33 | 34 | 35 | 36 | 37 | 38 | 39 | 40 | 41 | 42 | 43 | 44 | 45 | 46 | 47 | 48 | 49 | 50 | 51 | 52 | 53 |
| Closure/digital education | X | X | X | X | X | | | | | | | | | | | | | | | | | | | | | | | | | | | | | | | | | | | | | | | | | | | | | | | | |
| Half-occupancy (Hybrid education - 1/4 in-school) | | | | | | X | X | X | X | X | X | X | X | X | X | X | X | X | X | X | X | X | | | | | | | | | | | | | | | | | | | | | | | | | | | | | | | |
| Full in person | | | | | | | | | | | | | | | | | | | | | | | X | X | X | X | X | X | X | | | | | X | X | X | X | X | X | X | X | X | X | X | X | X | X | X | X | X | | | |
| 1.5 m distance to classmates | | | X | X | X | X | X | X | X | X | X | X | X | X | X | X | X | X | X | X | X | | | | | | | | | | | | | | | | | | | | | | | | | | | | | | | | |

| | | | | | | | | | | | | | | | | | | | | | | | | | | | | | | | | | | | | | | | | | | | | | | | | | | | | | |
|---|---|---|---|---|---|---|---|---|---|---|---|---|---|---|---|---|---|---|---|---|---|---|---|---|---|---|---|---|---|---|---|---|---|---|---|---|---|---|---|---|---|---|---|---|---|---|---|---|---|---|---|---|---|
| 1.5 m distance to teachers | | | X | X | X | X | X | X | X | X | X | X | X | X | X | X | X | X | X | X | X | X | X | X | X | X | X | X | X | | | | | X | X | X | X | X | X | X | X | X | X | X | X | X | X | X | X | X | | | |
| Antigen testing (2x pw preventive/risk-based) | | | | | | | | | | | | | | | | | X | X | X | X | X | X | X | X | X | X | X | X | X | | | | | X | X | X | X | X | X | X | X | X | X | X | X | X | X | X | X | X | | | |
| Masking | | | X | X | X | X | X | X | X | X | X | X | X | X | X | X | X | X | X | X | X | X | X | X | X | X | X | X | X | | | | | X | X | X | X | X | | | | | | | | | X | X | X | | | |
| Symptomatic persons quarantined | | | X | X | X | X | X | X | X | X | X | X | X | X | X | X | X | X | X | X | X | X | X | X | X | X | X | X | X | | | | | X | X | X | X | X | X | X | X | X | X | X | X | X | X | X | X | X | | | |
| Quarantine class (>2cases) | | | | | | X | X | X | X | X | X | X | X | X | X | X | X | X | X | X | X | X | X | X | X | X | X | X | X | | | | | X | X | X | X | X | X | X | X | X | X | X | X | X | X | X | X | X | | | |
| Holidays | | | | | | | | | | | | | | | | | | | | | | | | | | | | * | * | X | X | X | X | * | * | | | | | | | | | | | | | | | | X | X | X |
| *summer holidays differ per region. | | | | | | | | | | | | | | | | | | | | | | | | | | | | | | | | | | | | | | | | | | | | | | | | | | | | | |

Source: VO-Raad, 2022.[1]

**Appendix B. Detailed model description.**

**Agent-based model**

We developed an agent-based model to simulate SARS-CoV-2 transmission in a secondary school informed by data described in Appendix A. We give a detailed description below and present a summary of the model parameters in Table 1 in the main text of the manuscript. The code for the model can be found on Github.[2]

We distinguished two types of individuals: (1) students, characterised by the grade and class they belong to, (2) teachers, characterised by the classes they educate. Based on the average values reported in the pilot project (Appendix Table 1), the secondary school in our model comprises six grades with a varying number of classes and varying number of students per class per grade (Appendix Table 19). Consequently, the school encompasses 944 students in total. We further assumed that students attend five subjects per day and that teachers educate two to three classes per day, leading to 72 teachers in the model. In reality, the average workload of teachers in the Netherlands is 0.74-0.87 full-time equivalent per teacher, so teachers do not work full time at school and possibly do not teach at school every day.[3] In the model, we assumed teachers work every day but educated only two to three classes per day such that the working hours roughly match the average workload in the Netherlands.

**Appendix Table 19. School characteristics in the model.**

| Grade | Number of classes per grade | Number of students per class |
|---|---|---|
| 1 | 7 | 23 |
| 2 | 6 | 29 |
| 3 | 8 | 23 |
| 4 | 7 | 29 |
| 5 | 5 | 30 |
| 6 | 3 | 23 |

*Contact network*

We defined contacts relevant for transmission based on data from the pilot project (Appendix A.2), where students and teachers were surveyed about the number of people from their school they had a conversation with (at less than 1.5m distance and for at least 15 min), or had physically touched the day before. Students reported the number of contacts with other fellow students within the same class and outside their class during school hours and outside school-hours (Appendix Table 13 and 14). Results were translated into contact matrices for contacts between students and between teachers in the school environment (Appendix Table 20). Contacts between teachers and students were estimated from the number of students educated per teacher (Appendix Table 15), as these were not surveyed in a similar way.

We distinguished weekdays (Monday to Friday) and weekends (Saturday and Sunday), and divided the day into three periods of eight hours each, distinguished by the types of contacts:

1. *School hours:* Students and teachers have a certain number of within-school contacts described by the respective contact matrices (Appendix B).
2. *Outside school hours:* Students are assumed to have some school-related contacts during their leisure time activities. These contacts are randomly sampled from their within-school contacts (one within-class contact, one outside class contact). Teachers are assumed to have no contacts with other teachers after school hours. Transmission risks caused by school-unrelated contacts are modelled by a constant introduction rate of infected students and teachers (see *Infection risk from community*).
3. *Night hours:* Neither students nor teachers are assumed to have any contacts during this time.

Other types of contacts (e.g., household contacts) are not implemented in our model. Infections from other types of contacts are treated as importations into the school (see *Infection risk from community*).

*Contacts between students*

The contact matrix representing the number of contacts relevant for transmission that a student has with other students from different grades, or with other students within his/her own class is based on the mean reported values from the survey (Appendix Table 17):

**Appendix Table 20. Number of contacts between students of different grades.** The first six columns represent the number of contacts a student of the respective grade have outside of their class but within school and the last column represents the number of contacts within their own class.

| Grade | 1 | 2 | 3 | 4 | 5 | 6 | Within-class |
|---|---|---|---|---|---|---|---|
| **1** | 3 | 0 | 0 | 0 | 0 | 0 | 6 |
| **2** | 2 | 5 | 0 | 0 | 0 | 0 | 5 |
| **3** | 1 | 1 | 5 | 0 | 0 | 0 | 6 |
| **4** | 1 | 1 | 2 | 7 | 1 | 1 | 8 |
| **5** | 0 | 0 | 0 | 1 | 7 | 1 | 8 |
| **6** | 0 | 0 | 0 | 1 | 2 | 7 | 11 |

Based on data from the pilot project (Appendix Table 16), students are assumed to have one close contact within their class (i.e., conversation within 1.5m distance for > 15 minutes or physical touch) and one close contact outside of their class but within the school. These close contacts are eligible for quarantine if the student is infected and develops symptoms.

Outside of school hours students are assumed to meet two other students (pilot project, Appendix Table 13). These contacts are randomly sampled from their within-school contacts.

*Contacts between teachers*

Based on the median number of contacts reported in the pilot project, we used six contacts between teachers during school hours (Appendix Table 15) and no contacts between teachers outside school hours (Appendix Table 13). These contacts were randomly sampled each day.

*Contacts between teachers and students*

We assumed that contacts relevant for transmission only occur with a proportion of all students a teacher educates per day, based on close conversations or proximity to the teacher. In our model, the effective number of transmission-relevant contacts between teachers and students varies between 8 and 10 (median number reported in Appendix Table 6). Since typically, teacher-student contacts are not as close as contacts between students, we assumed that contacts between teachers and students have a reduced probability of transmission relative to contacts between students or contacts between teachers. The reduced probability of transmission is uniformly distributed with a minimum of 15% and a maximum of 50%.

*Transmission model*

Individuals may be either susceptible, vaccinated, symptomatically infected, asymptomatically infected, or recovered. We assumed that students have on average a 46% reduced susceptibility to infection when compared to teachers, based on estimates from studies on earlier variants.[4] Symptomatically infected individuals are assumed to develop symptoms according to a Weibull-distributed incubation period (mean = 2.7 days, sd=1.74), assuming a 60% mean incubation period compared to estimates for the wild-type virus.[5]

*Infectivity*

Infected individuals are assumed to be readily infectious with a time-varying level of infectivity. Infectivity is assumed to be on average 50% lower for asymptomatic than for symptomatic individuals, and 15% lower for students when compared to teachers.[6] The baseline infectivity is distributed according a Gamma distribution (mean = 2.2 days) based on results specific for Omicron[7] and the respective reproduction number (average number of secondary cases caused by an infectious individual):

We denote infectivity over time since infection $\tau$ by $\beta(\tau)$. It is the mean rate at which an individual infects others at time $\tau$ after its time of infection. We use the infectivity profile for calculating the probability of transmission from an infectious to a susceptible individual (see below). The reproduction number R is given by integrating $\beta(\tau)$ over time since infection $R=\int \beta(\tau)d\tau$. The generation time distribution $\omega(\tau)$ is given by unit normalisation such that $\omega(\tau)=\beta(\tau)/R$. Assuming the mean generation time to be equivalent with the observed mean serial interval, we calculate the infectivity profile by $\beta(\tau)=\omega(\tau)R$. We assumed that infectivity over time since infection differs between asymptomatic and symptomatic infected individuals (Table 1 in the main text).

*Asymptomatic infections*

The proportion of asymptomatic infections is assumed to be different for students and for teachers. We sampled the probability of developing an asymptomatic infection from a Uniform(0.17, 0.25) and Uniform(0.15, 0.6) distribution for teachers and students, respectively. For students, these values are based on data reported in the pilot project and are in agreement with estimates by Buitrago-Garcia and colleagues.[8] For teachers, we used the overall estimate reported in Buitrago-Garcia and colleagues.[8]

*Aerosol transmission*
Additionally to transmission through direct contacts, we assume that students and teachers may become infected through aerosol transmission: For each infected student or teacher, susceptible students or teachers in the same classroom may become infected by assuming a transmission probability equivalent to a direct contact with the index case. However, we instigate aerosol transmission only in 10% of the potential transmission events, representing superspreading events in a classroom.

*Seasonality*
We distinguished a winter period (October till March) and summer period (April till September) and assumed a 25% decrease in the reproduction number during the summer season.

*Infection risk from community*
We assumed that all susceptible individuals are exposed to a certain school-unrelated risk of infection. For simplicity, we assumed a constant probability of infection from community (school-unrelated) each week. The probabilities are based on an age-dependent community incidence between 180-300/100.000 per week in October 2021.[9] Quarantined individuals are assumed to be excluded from this exposure.

*Holidays*
We assumed no within-school transmission during holidays. Since holidays may differ between countries, we implemented only summer and Christmas holidays (15th July – 1st September and 15th December – 3rd January, respectively). Students and teachers may become infected from school-unrelated contacts in the community (see *Infection risk from community*).

*Accuracy of the diagnostic test*
In our model, compliant symptomatically infected students are assumed to be tested using reverse transcriptase polymerase chain reaction (RT-PCR). These students are treated as symptomatic index cases and may trigger the quarantine of all classmates and close contacts in the quarantine intervention scenario. These contacts may exit quarantine on day five after the start of quarantine if they are tested negative using a rapid antigen test. For screening interventions, students and teachers are also tested using a rapid antigen test. We assumed a time-varying imperfect sensitivity for both diagnostic tests (Figure 1 in the main text) based on results reported in Smith and colleagues and adjusted for a shorter generation time for the Omicron variant.[7,10] Throughout the simulations, we assume the test sensitivity to be the same for symptomatic and asymptomatic infections, and we assume a specificity of 100% for both PCR and antigen tests.

*Vaccination*
Based on data from the Netherlands, we assumed a vaccination coverage of 60% for students and 80% for teachers.[11] Vaccinated teachers are assumed to have received one booster shot. No booster doses were assumed for students. Vaccine efficacies in reducing susceptibility to infection are based on efficacies reported for previous variants and then scaled by a factor, representing reduced efficacy for the Omicron variant (Table 1 in Keeling and colleagues).[12,13] We assumed no difference between infectivity of breakthrough infections in vaccinated and non-vaccinated individuals.[14] Vaccination times are assigned according to data from the Netherlands.[11] We assumed no direct effect of vaccination on infectivity but it reduces the probability of developing a symptomatic infection, thereby indirectly lowering infectivity.

*Timing of previous infection*
We loosely based the proportion of students and teachers infected by SARS-CoV-2 prior to the study period on an antibody study performed by Sanquin in the Netherlands.[15] We assumed a seroprevalence of 35% for students and 30% for teachers. The respective infection times were based on the SARS-CoV-2 incidence in the Netherlands since the beginning of the epidemic in February 2020.[16]

*Waning of immunity*
We allowed for reinfections after recovery from natural infection or after vaccination. Sterilising immunity wanes according to an exponential decline with an average time of waning of nine months and was informed by Townsend and colleagues (Appendix Figure 1A).[17] The authors estimated the antibody waning profile over time for SARS-CoV-2 for IgG antibody levels to the spike protein, nucleocapsid protein, and to the whole viral lysate using phylogenetic analysis of the ancestral and descendent states. We assumed a direct correspondence between the antibody decline and the waning of sterilising immunity of infected or vaccinated individuals. We implemented an all-or-nothing waning function, where individuals return to the susceptible state with a certain probability each day derived from the corresponding antibody waning profile. In our baseline scenario, sterilising immunity wanes

equally for individuals that recover from natural infection and those that were vaccinated. We further assumed that individuals return to only 75% of their original susceptibility value, representing residual protection from previous exposure to the virus. At each reinfection, the proportion of symptomatic infections is assumed to be reduced by 20%.

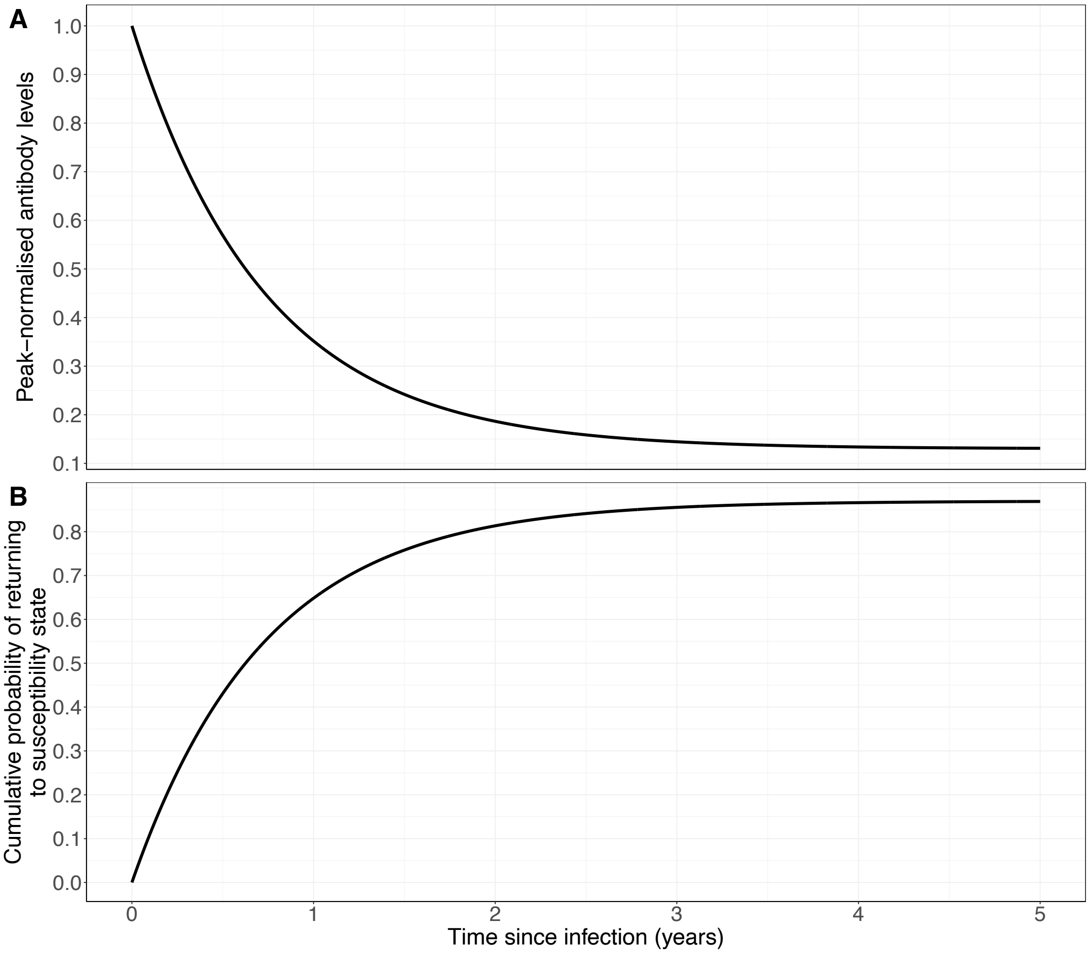

**Appendix Figure 1. Antibody waning profile and probability of susceptibility used in the model.** (A) Peak-normalised coronavirus anti-spike protein IgG antibody levels over time for SARS-CoV-2. The antibody waning profile is based on Townsend and colleagues.[17] (B) Cumulative probability of returning to the susceptible state after being infected with of vaccinated against SARS-CoV-2.

**Simulation scenarios**

Simulations were performed over a course of 30 months assuming a start date of 3rd January 2022. We explored the effect of variation of several parameters on the transmission dynamics and on the percentage of students who are (symptomatically) infected, susceptible, and absent due to isolation and (if applicable) quarantine for each week of the study period. Absenteeism among students per week is calculated as the number of students that are absent at least one day during the respective week. The number of susceptible students is weighed according to their susceptibility value (baseline susceptibility to infection of 100% is represented by unvaccinated teachers). We defined the health burden on students as the number of symptomatic student days and computed the reduction for each scenario compared to the baseline scenario. Since the annual booster campaign is only in effect after 1$^{st}$ September 2022, we computed the health burden for the intervention scenarios starting from that date. As a cost-benefit measure for intervention scenarios, we computed the number of prevented infections per absent student. Fixed parameters and those used in the baseline scenario are given in Table 1 of the main text. Parameters that are varied in other scenarios are given in Table 2 of the main text.

*Baseline scenario*

The baseline scenario assumes a school-related reproduction number for Omicron of 2.0 during the winter period (October to March) and of 1.5 during the summer period (25% decrease compared to winter, April to September), assuming a 40%-100% increase to estimates from the Delta variant.[18,19] We assumed compliance to isolation guidelines for symptomatically infected students of 33%, i.e. home isolation for seven days upon a positive PCR test, but no other mitigation measures in schools such as quarantine of exposed contacts, screening policies, mask mandates, or class size reductions.[20]

*Scenario: Varying reproduction number.*
We distinguished two scenarios to account for the uncertainty in the school-related reproduction number: (a) 50% lower reproduction number in winter ($R_{winter}$ = 1.0, $R_{summer}$ = 0.75), (b) 100% higher reproduction number in winter ($R_{winter}$ = 4.0, $R_{summer}$ = 3.0).

*Scenario: Susceptibility to reinfection*
We distinguished (a) a lower susceptibility to reinfection of 50% and (b) full susceptibility to reinfection, i.e., 100% of the original susceptibility value.

*Scenario: Vaccine immune escape*
We assumed (a) 25% lower and (b) 25% higher average vaccine efficacy in reducing susceptibility to reinfection, reflecting higher and lower immune escape in vaccinated individuals, respectively.

*Scenario: Waning of immunity*
We investigated two alternative average durations of sterilising immunity: (a) 3 months and (b) 18 months, as opposed to 9 months for the baseline scenario.

*Intervention scenario: Quarantine of close contacts and classmates*
Upon a positive test result of a compliant symptomatically infected student, all close contacts and classmates quarantine for ten days. We assumed that teachers do not have to quarantine. The quarantine period may be shortened if the individual has a negative antigen test result on day five after the start of quarantine. We assumed an increased case detection in comparison with the baseline scenario and distinguished (a) 50% and (b) 75% symptomatic case isolation.

*Intervention scenario: Regular screening*
A proportion of students will perform an antigen test twice weekly with (a) 50% and (b) 75% adherence to this screening intervention.

*Intervention scenario: Annual booster of students and teachers*
All students and teachers that were fully vaccinated are assumed to receive one booster vaccination dosis during each summer holidays. Vaccine efficacy reducing the susceptibility to infection for booster vaccination is equal to the initial efficacy increased by 20% (minimum and maximum value of the distribution is increased by 20%).

*Sensitivity analyses*
We performed the following sensitivity analyses:

- Higher symptomatic case isolation (90%)
- Equal infectivity of asymptomatically and symptomatically infected individuals
- No increase in vaccine efficacy after booster vaccination
- Different waning rates after natural infection vs vaccination

More details and results can be found in Appendix E.

**Appendix C. Implementation of the model.**

The model was built using R (version 4.0.1). The code is available from [ref]. A description of the most important processes of the agent-based model are given below.

**Study period and scheduling**
The simulation is run for 868 days (30x4 weeks) in total. Events take place at three time points each day, representing school hours, leisure hours, and night hours. All presented scenarios are run for 100 simulations.

**Initialization**

At the beginning of each simulations the following events take place in the following order:

1. *Set up grades and classes:* Go through all grades and classes according to Appendix Table XX and assign the corresponding number of students to a class.
2. *Set up contact network:* Go through all students individually and randomly assign a contact to students from the same class or from other classes according to the contact matrix in Appendix Table 20. Contacts are made symmetric by automatically assigning the contact to the randomly drawn student as well. A similar procedure is performed for contacts outside school and for contacts between teachers.
3. *Assign teachers to classes:* Teachers are randomly assigned to either teaching subject 1 and 2 to grades 1-3 or 4-6, or subjects 3-5 to grades 1-3 or 4-6.
4. *Importations from community:* Susceptible students and teachers are randomly chosen to be infected in the community. The number of infected individuals from community is ensured to meet a fixed number.
5. *Vaccination prior to study period:* Students and teachers are randomised (according to the respective vaccination coverage) to either being vaccinated or unvaccinated at the beginning of each simulation.
6. *Susceptibility values:* Unvaccinated teachers are assumed to be fully susceptible (susceptibility value is set to one). Vaccinated teachers have a reduced susceptibility to infection based on the assumed vaccine efficacy distribution (Table 1 in main text). Students are assumed to have a reduced susceptibility based on the relative susceptibility distribution assumed in the model (Table 1 in main text). Their susceptibility is further reduced if students are vaccinated (according to the vaccine efficacy distribution in Table 1 in the main text). Recovered individuals are assumed to be have a susceptibility value of zero.
7. *Previous infection times:* For each individual that was randomised to have been infected before the study period, an infection time was drawn according the infection data of the Netherlands.[16]
8. *Initial reinfections:* Based on the assumed daily probability of waning of immunity, previously infected individuals may be reinfected at the beginning of the study period.
9. *Screening adherence:* Based on the assumed screening adherence, students are randomized to participate in screening.
10. *Compliance to isolation:* Based on the assumed proportion of symptomatic case isolation (Table 1 in main text), individuals are randomized to comply with isolation rules.
11. *Compliance to quarantine:* Based on the assumed proportion of quarantine compliance (Table 1 in main text), individuals are randomized to adhere with quarantine rules. We assumed no correlation between individuals that comply to isolation and those that comply to quarantine rules.

**Events in each simulation**

In our simulations, events can take place at three time steps within one day: during school hours ($t_s$), outside school hours ($t_o$), and during night hours ($t_n$).
The following events take place at certain time steps (which one is indicated in the description) of the simulation in the following order:

1. *Teacher-Student contacts:* Sample contacts between teachers and students at the beginning of each day ($t_s$).
2. *Booster vaccination of students and teachers:* On the first day of school after each summer holiday, a new vaccination time is randomly drawn from the time period of the previous summer holiday for each student and teacher. The respective individual is susceptible again (retrospective to the new vaccination date). The susceptibility value is determined according to a vaccine efficacy that was increased by 50%.
3. *Introductions from community:* Randomly infect susceptible students and teachers according to the fixed probability of infection from the community at $t_o$. Update the susceptibility status of newly infected individuals.
4. *Isolation of symptomatically infected:* Isolate all students and teachers at the beginning of each day ($t_s$).
5. *Quarantine:* Quarantine classmates and close contacts of symptomatic index cases at the beginning of each day ($\mathrm{t_s}$).

6. *Release students from quarantine:* For each quarantined student, perform an antigen test on day 5 of quarantine and release the student from quarantine if respective test result is negative.
7. *Reinfections:* At each time step of the day, students and teachers may become susceptible to reinfection. Waning is implemented as "all-or-nothing" using a Bernoulli trial and the waning of immunity probability distribution. If a recovered individual becomes susceptible again, the
8. *Screening intervention:* Twice-weekly screening is performed on Mondays and Wednesdays at the beginning of the day before any other events like transmission take place. Every time the screening function is called, individuals eligible for testing, i.e., those who are not isolated and adhere to screening are testing using the sensitivity of an antigen test (Figure 1D in the main text). We assumed no correlation between vaccination status and adherence to screening. Individuals who participate in screening are randomly chosen (according to the pre-set screening adherence) at the beginning each simulation. In addition, we assumed that individuals who participate in screening also adhere to isolation rules upon a positive test.
9. *Transmission:* Transmission events between students and teachers may take place during or outside school hours. For all currently infected individuals, transmission events to their susceptible contacts are performed. This may depend on the specific time during the day and whether it is currently weekday or weekend. For transmissions in class rooms, we allow for transmission from direct contacts and for aerosol transmission (see description *Aerosol transmission* in Appendix B). Infected individuals are immediately immune to infection (so non-susceptible), but their immunity may wane as described in "6. Reinfections").

**Appendix D. Additional figures and results.**

We present below additional figures supplementing the analyses shown in the main text of the manuscript.

**Data and model results comparison**

We compared the model results from the baseline scenario of our model to data on the incidence in children and young adults in the age group 10-19 in the Netherlands (Appendix Figure 2). Both the data and the model results show a large outbreak at the beginning of 2022 and low levels of infections by the end of March 2022. Our model results show a peak in newly infected students in the model school in the week of 10th January 2022 (Appendix Figure 2B). The Dutch data on newly positive tested individuals show a peak in the week of 24th January 2022. Note that we would expect a delay in positive tests (with respect to onset of infections) and that our results show predictions only for one school while the data is aggregated for the whole country of the Netherlands.

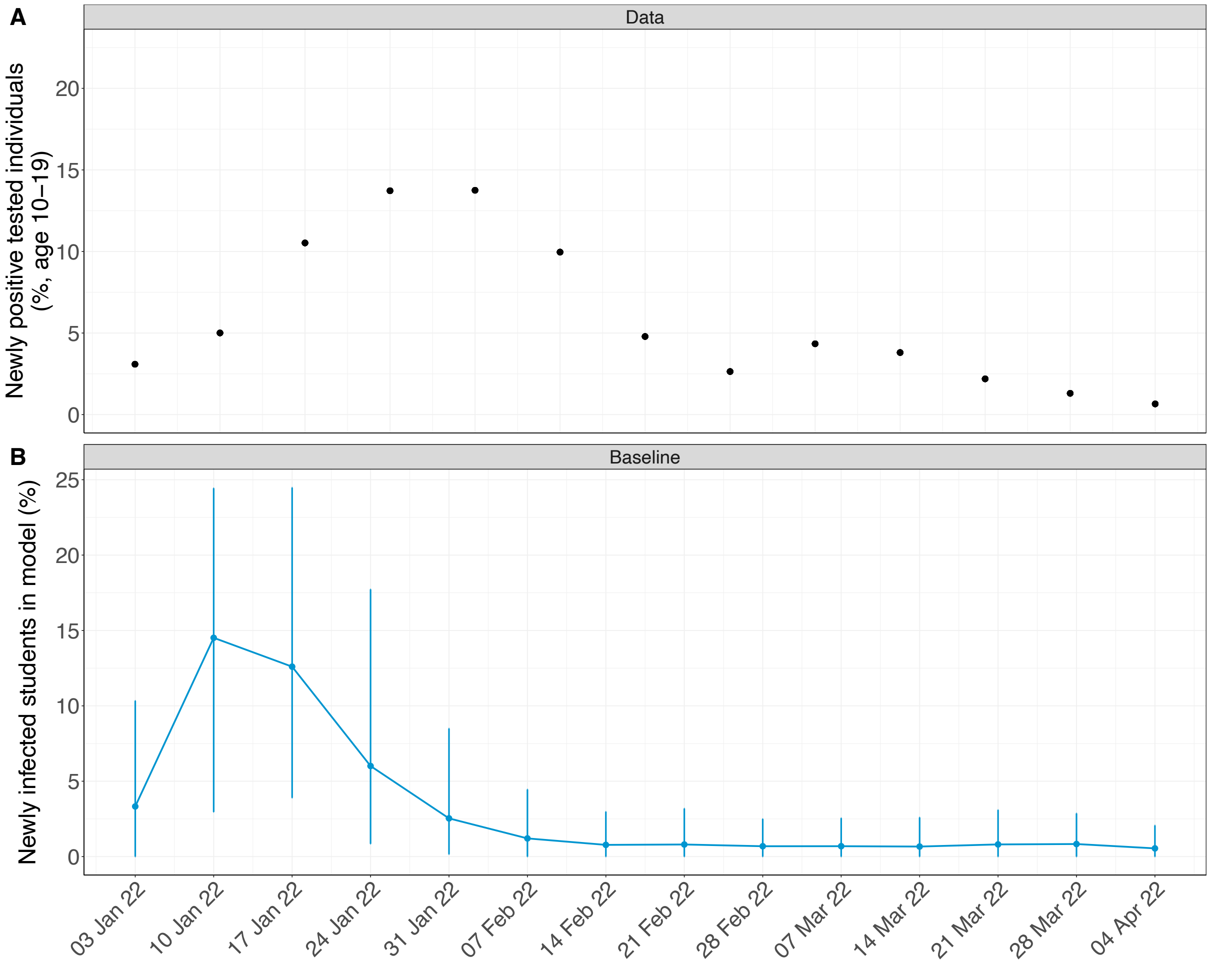

**Appendix Figure 2. Comparison of model results and data on positive tested 10-19-year-olds in the Netherlands in January till April 2022.** (A) Percentage of individuals aged 10-19 who were newly tested positive from 3rd January till 4th April 2022 per week. (B) Percentage of newly infected students as predicted from the baseline scenario from 3rd January till 4th April 2022 per week.

### Total number of symptomatic student days

The plot below shows the total number of symptomatic student days for each simulation scenario. Figure 7 in the main text is computed with respect to the total number of symptomatic student days as predicted in the baseline scenario. Similar to the main conclusions, the plot shows that the duration waning of immunity has the largest impact on the outcome. Among the interventions, screening is most effective in reducing the number of symptomatic student days. Note, however, that the effect of the annual booster campaign becomes similar to screening if it is evaluated over the relevant time period, i.e., after the boosters have been administered (as shown in the main text).

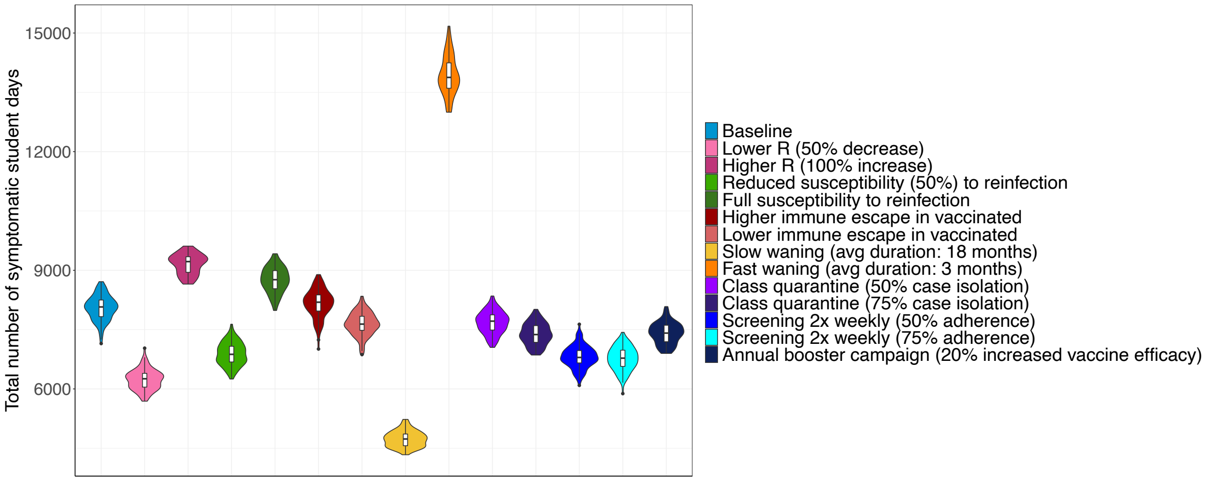

**Appendix Figure 3. Total number of symptomatic student days for the simulation scenarios.** The sum of days that students are in a symptomatic state during the whole study period (03/01/2022 till 24/05/2024) is shown for all simulation scenarios.

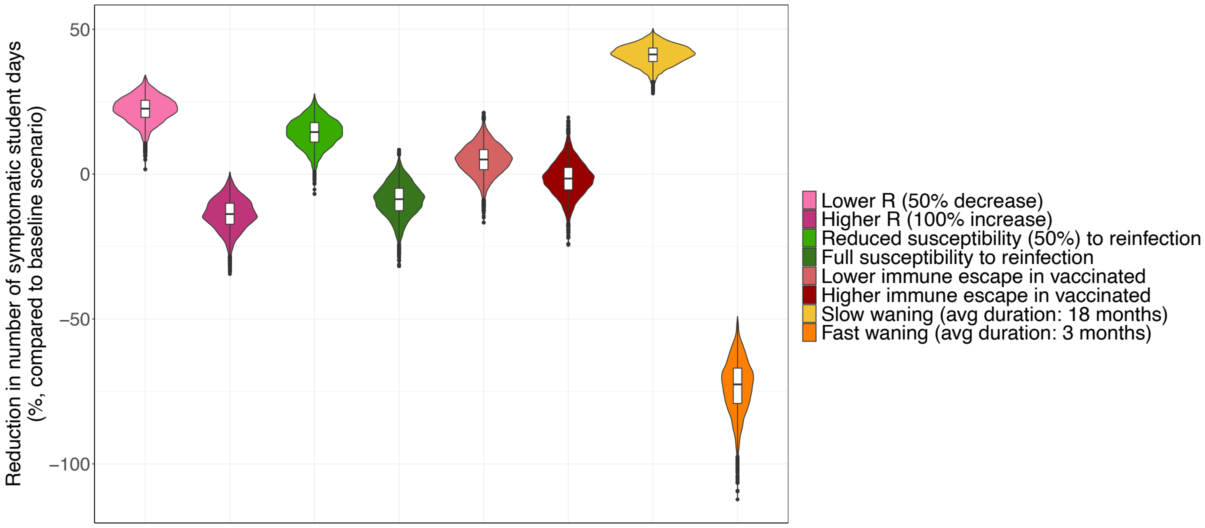

**Appendix Figure 4. Reduction in number of symptomatic student days for simulation scenarios.** The health burden (reduction in total number of days that students are in a symptomatic state) during the whole study period (03/01/2022 till 24/05/2024) is shown for all simulation scenarios.

### Symptomatically infected students

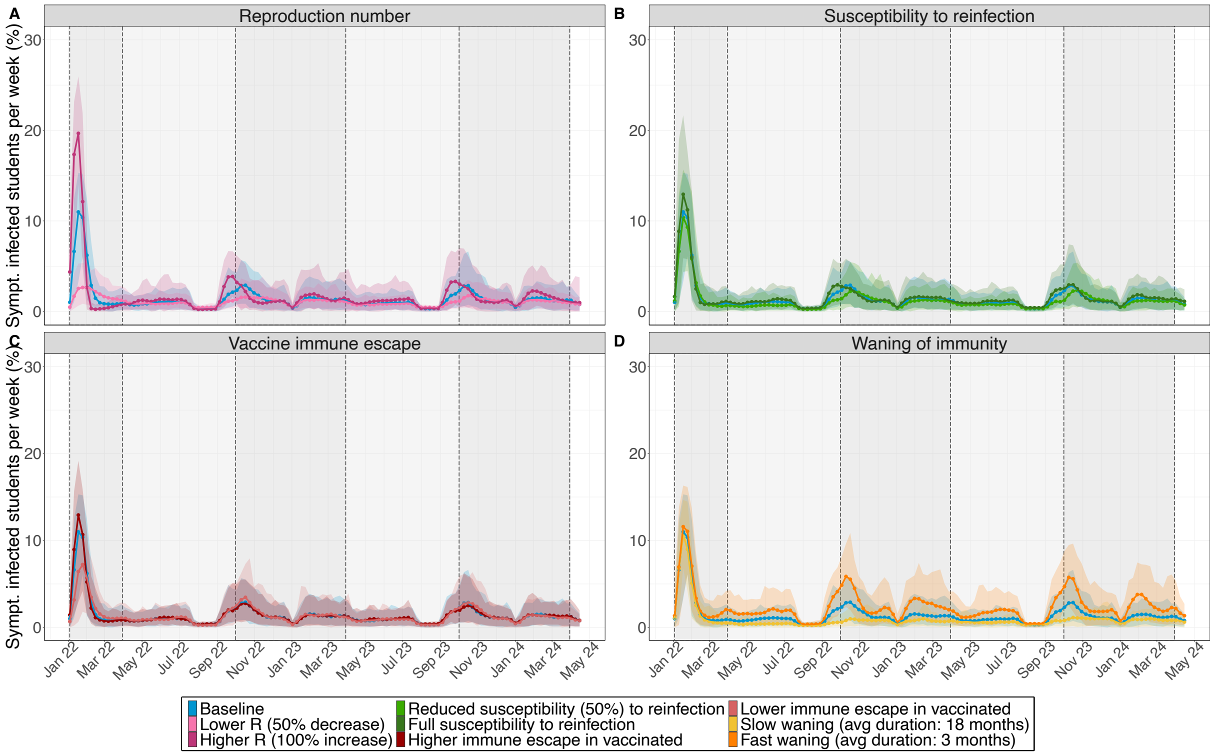

**Appendix Figure 5. Symptomatically infected students for the simulation scenarios.** The percentage of symptomatically infected students is shown for each week from 03/01/2022 till 24/05/2024. Bold points represent the mean value per week over 100 simulations. Shaded coloured areas are 95% uncertainty intervals over 100 simulations. Dark grey background represents the "winter" period (October till March). Light grey background represents the "summer" period (April till September). (A) Scenarios where the within-school reproduction number is varied: (i) lower ($R_{winter} = 1.0, R_{summer} = 0.8$) and (ii) higher reproduction number ($R_{winter} = 4.0, R_{summer} = 3.2$). (B) Scenarios where susceptibility to reinfection is varied: (i) susceptibility to reinfection is reduced by 50% of the original susceptibility value and (ii) the full susceptibility to reinfection. (C) Scenario with lower vaccine efficacy against susceptibility of infection. (D) Scenarios where average duration of waning of immunity is varied: (i) slow waning (average duration: 18 months) and (ii) fast waning (average duration: 3 months).

**Absent students**

Generally, the dynamics of absenteeism among students follows the dynamics of infected students (Figure 3 in the main text).

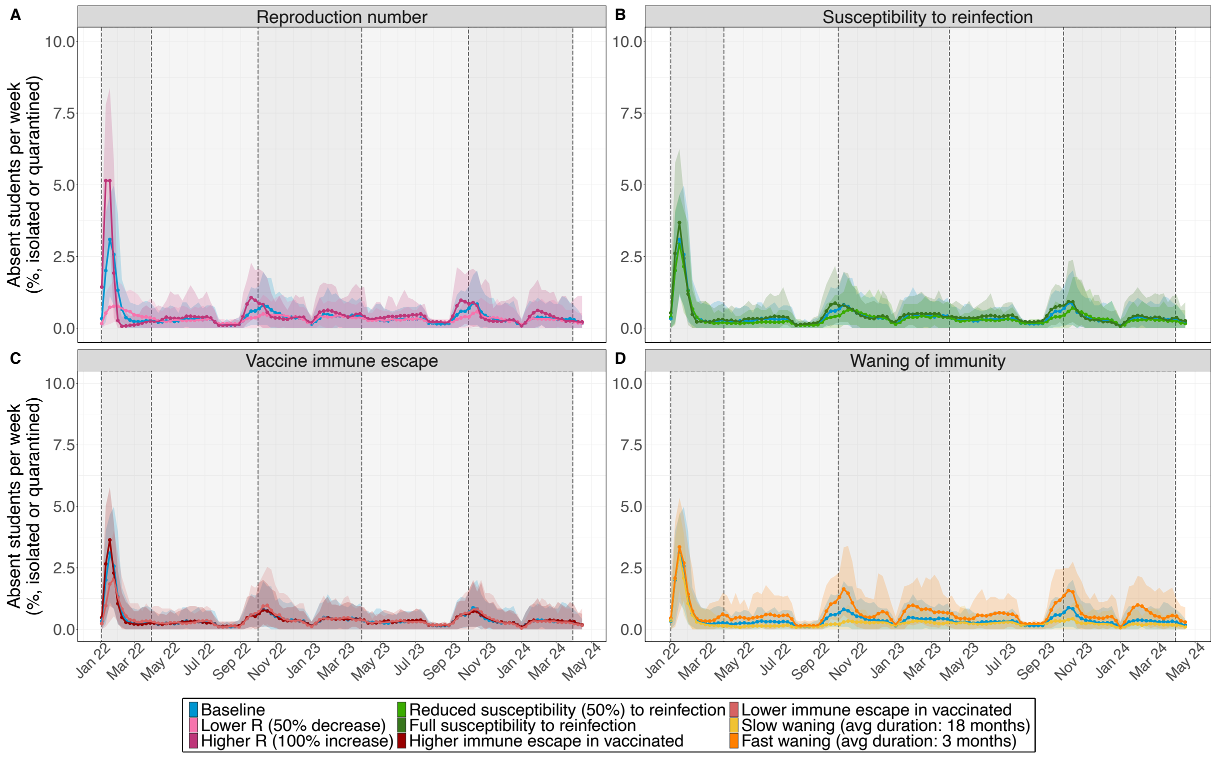

**Appendix Figure 6. Absenteeism among students for the simulation scenarios.** The percentage of absent students due to quarantine or isolation is shown for each week from 03/01/2022 till 24/05/2024. Bold points represent the mean value per week over 100 simulations. Shaded coloured areas are 95% uncertainty intervals over 100 simulations. Dark grey background represents the "winter" period (October till March). Light grey background represents the "summer" period (April till September). (A) Scenarios where the within-school reproduction number is varied: (i) lower ($R_{winter} = 1.0, R_{summer} = 0.8$) and (ii) higher reproduction number ($R_{winter} = 4.0, R_{summer} = 3.2$). (B) Scenarios where susceptibility to reinfection is varied: (i) susceptibility to reinfection is reduced by 50% of the original susceptibility value and (ii) the full susceptibility to reinfection. (C) Scenario with lower vaccine efficacy against susceptibility of infection. (D) Scenarios where average duration of waning of immunity is varied: (i) slow waning (average duration: 18 months) and (ii) fast waning (average duration: 3 months).

**Weighted susceptibility to infection**

We assumed that unvaccinated teachers have a susceptibility of 100% for infection. The number of students at risk for infection is weighted according to their susceptibility value which is reduced due to an assumed reduced susceptibility of students when compared to teachers and further reduced if the student is vaccinated.

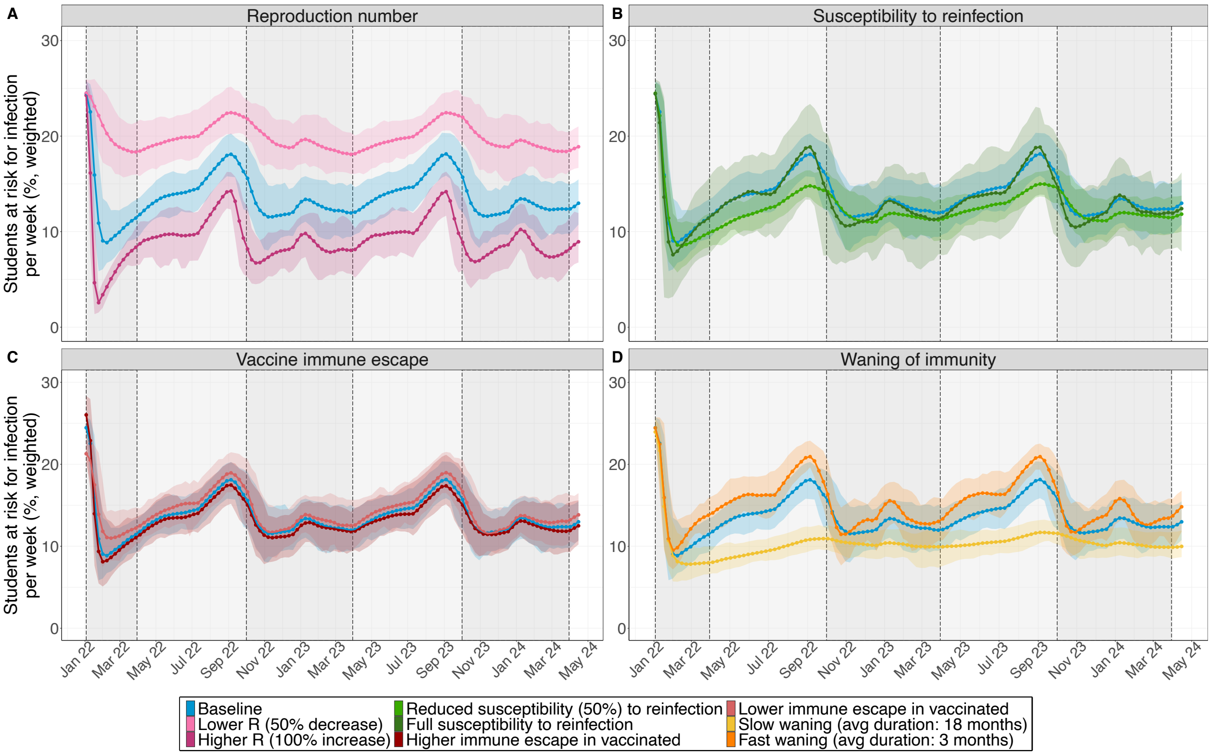

**Appendix Figure 7. Students at risk for infection for the simulation scenarios.** The percentage of students at risk for infection is shown for each week from 03/01/2022 till 24/05/2024. The number of students at risk for infection is weighted according to their susceptibility value (with a baseline of 100% for unvaccinated teachers). Bold points represent the mean value per week over 100 simulations. Shaded coloured areas are 95% uncertainty intervals over 100 simulations. Dark grey background represents the "winter" period (October till March). Light grey background represents the "summer" period (April till September). (A) Scenarios where the within-school reproduction number is varied: (i) lower ($R_{winter} = 1.0, R_{summer} = 0.8$) and (ii) higher reproduction number ($R_{winter} = 4.0, R_{summer} = 3.2$). (B) Scenarios where susceptibility to reinfection is varied: (i) susceptibility to reinfection is reduced by 50% of the original susceptibility value and (ii) the full susceptibility to reinfection. (C) Scenario with lower vaccine efficacy against susceptibility of infection. (D) Scenarios where average duration of waning of immunity is varied: (i) slow waning (average duration: 18 months) and (ii) fast waning (average duration: 3 months).

**Appendix E. Sensitivity analyses.**

We evaluated the changes of our results with respect to changes in our model parameters. We present the results and corresponding plots below.

**Higher proportion of symptomatic case isolation**

We performed a sensitivity analysis to test the impact of a 90% symptomatic case isolation on our results (Appendix Figure 8). General dynamics (number of wave) are preserved when more symptomatic cases are isolated. However, the sizes of the outbreaks are much smaller and the peaks occur a bit later when compared to the baseline scenario in the main text of the manuscript. Expectedly, the peaks of the percentage of absent students is tripled.

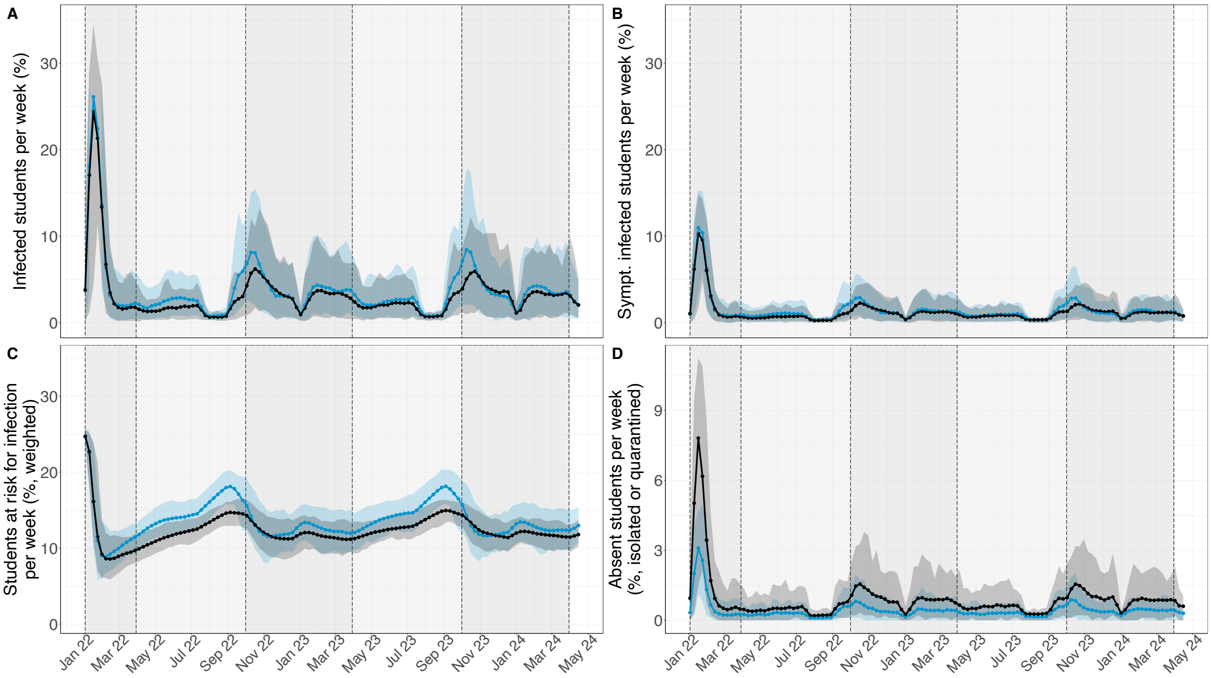

**Appendix Figure 8. SARS-CoV-2 transmission for baseline scenario (blue) and scenario with 90% symptomatic case isolation (black). Numbers are given for each week in the study period.** (A) Proportion of students infected due to school-related infections and introductions from community. (B) Proportion of students symptomatically infected per week. (C) Proportion of students at risk for infection per week, weighted by their susceptibility value (that depends on their vaccination status and the corresponding efficacy). (D) Proportion of students either isolated or quarantined per week. Bold points represent the mean value per week over 100 simulations. Shaded coloured areas are 95% uncertainty intervals over 100 simulations. Dark grey background represents the "winter" period (October till March). Light grey background represents the "summer" period (April till September).

### Equal infectivity of asymptomatically and symptomatically infected individuals

In a sensitivity analysis, we tested the impact of assuming equal infectivity of asymptomatically and symptomatically infected individuals on our results (Appendix Figure 9). Mainly the first wave after the Christmas holidays 2021 is affected with a higher peak in the scenario when asymptomatically and symptomatically infected individuals are equally infectious. This can be explained by the fact that the probability of developing a symptomatic infection is decreased by 20% after each reinfection in our model. This assumption becomes, thus, less important the more asymptomatic infections occur.

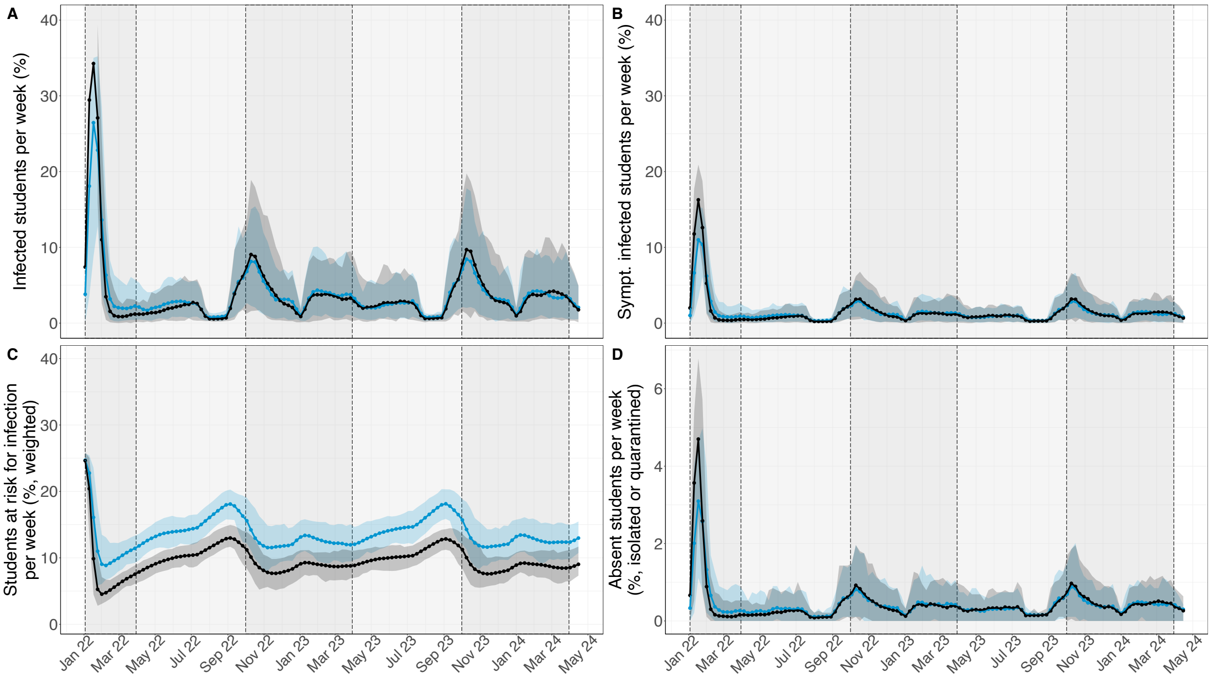

**Appendix Figure 9. SARS-CoV-2 transmission dynamics for the baseline scenario and a scenario (blue) with equal infectivity of asymptomatic and asymptomatic infections (black).** (A) Proportion of students infected due to school-related infections and introductions from community. (B) Proportion of students symptomatically infected per week. (C) Proportion of students at risk for infection per week, weighted by their susceptibility value (that depends on their vaccination status and the corresponding efficacy). (D) Proportion of students either isolated or quarantined per week. Bold points represent the mean value per week over 100 simulations. Shaded coloured areas are 95% uncertainty intervals over 100 simulations. Dark grey background represents the “winter” period (October till March). Light grey background represents the “summer” period (April till September).

### Differential waning of sterilising immunity for natural infection vs vaccination

We tested the impact of a faster duration of waning of sterilising after vaccination (3 months) than after a natural infection (9 months). This assumption mainly effects the first wave and transition to the summer in 2022, and the autumn wave in 2022 (Appendix Figure 10). A slightly higher first peak in January 2022 would be expected as well as a small resurgence in April 2022. The autumn peak is also expected to be larger. The general transmission dynamics are, however, similar to the baseline scenario.

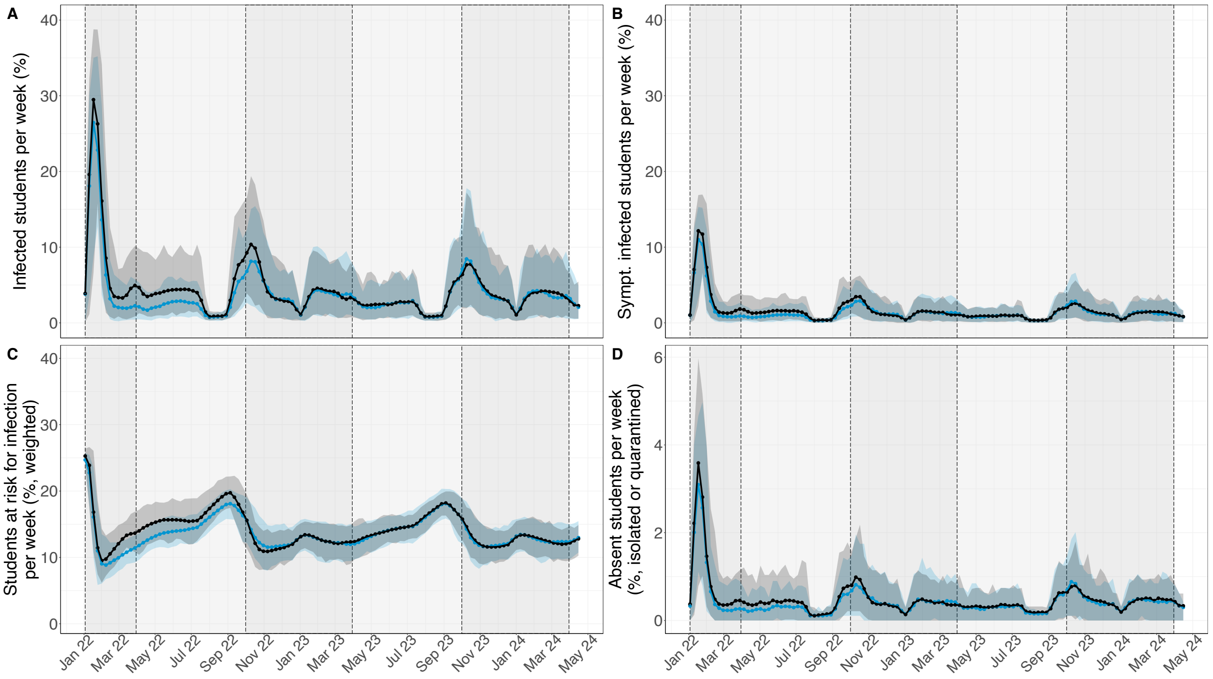

**Appendix Figure 10. SARS-CoV-2 transmission dynamics for the baseline scenario (blue) and a scenario with faster waning after vaccination vs natural infection (black).** In the baseline scenario sterilising immunity wanes on average after nine months. In the sensitivity scenario, immunity wanes on average after three months for vaccinated individuals and after nine months for naturally infected individuals. (A) Proportion of students infected due to school-related infections and introductions from community. (B) Proportion of students symptomatically infected per week. (C) Proportion of students at risk for infection per week, weighted by their susceptibility value (that depends on their vaccination status and the corresponding efficacy). (D) Proportion of students either isolated or quarantined per week. Bold points represent the mean value per week over 100 simulations. Shaded coloured areas are 95% uncertainty intervals over 100 simulations. Dark grey background represents the "winter" period (October till March). Light grey background represents the "summer" period (April till September).

#### Annual booster vaccinations for students and teachers

All students and teachers who were fully vaccinated are assumed to receive one booster dosis during the summer holidays each year. We varied the assumed vaccine efficacy after a booster vaccination: (1) The same vaccine efficacy as the initial vaccine efficacy used in the baseline scenario (Table 1 in main text), (2) 50% increase in vaccine efficacy for vaccinated individuals with a booster dosis, i.e., vaccine efficacy for students: Uniform(0.825, 0.855) and for teachers: Uniform(0.6, 0.885). Appendix Figure 11 shows that the effect of annual booster campaigns strongly depends on the assumed vaccine efficacy.

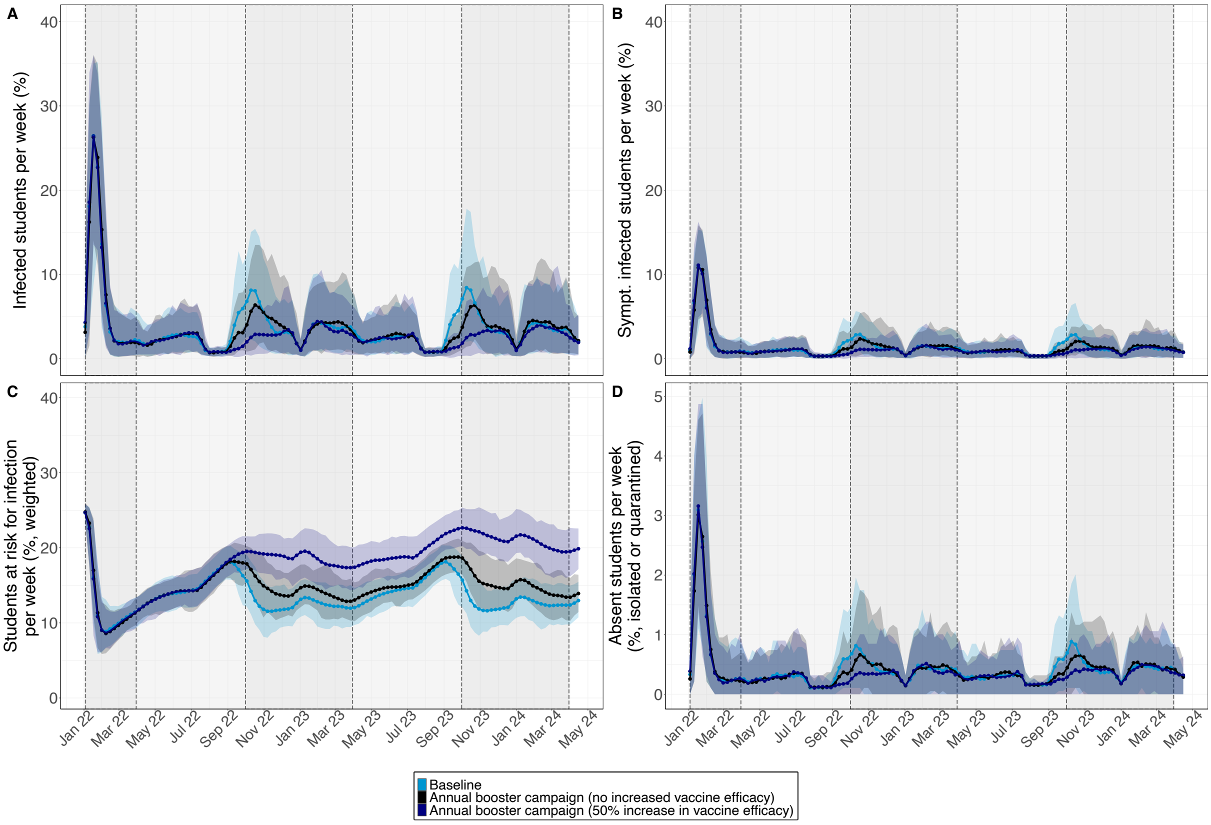

**Appendix Figure 11. SARS-CoV-2 transmission dynamics for annual booster vaccination of students and teachers.** All students and teachers who were fully vaccinated are assumed to receive one booster dosis during the summer holidays each year. Vaccine efficacies (VE) after booster vaccination is varied. Black: No increase in VE. Navy blue: 50% increase for booster vaccinations with respect to VE in the baseline scenario. (A) Proportion of students infected due to school-related infections and introductions from community. (B) Proportion of students symptomatically infected per week. (C) Proportion of students at risk for infection per week, weighted by their susceptibility value (that depends on their vaccination status and the corresponding efficacy). (D) Proportion of students either isolated or quarantined per week. Bold points represent the mean value per week over 100 simulations. Shaded coloured areas are 95% uncertainty intervals over 100 simulations. Dark grey background represents the "winter" period (October till March). Light grey background represents the "summer" period (April till September).
